# Whole exome sequencing reveals UNC45B as a novel candidate gene functionally associated with Dilated Cardiomyopathy

**DOI:** 10.64898/2025.11.28.25340252

**Authors:** Amrita Mukhopadhyay, Dharmendra Jain, Ashok Kumar, Bhagyalaxmi Mohapatra

## Abstract

UNC45B, a UCS class myosin chaperone that facilitates Hsp90-myosin interactions, plays a crucial role in early myofibrillogenesis, though its specific function sustaining cardiac contractility remains obscure. Here we identified seven non-synonymous (p.L486F, p.C487Y, p.A492T, p.D496Y, p.R721Q, p.A780V and p.C786Y) and two synonymous (p.Y769= and p.A780=) variants in *UNC45B* through whole-exome sequencing (WES) and subsequent genetic screening of Dilated Cardiomyopathy (DCM) patients by Sanger sequencing. The majority of the variants (77.7%) are localized within the highly conserved UCS domain of UNC45B, a critical region for chaperone-like property, also known for myosin binding site*. In-vitro* analysis by immunofluorescence revealed cytoplasmic mislocalization of mutant UNC45B and pronounced hypertrophic morphology, most evident in p.R721Q and p.A780V mutants. Nuclear fragmentation, a hallmark of cellular stress and apoptosis, was also prominently detected in these variants. Complementary ultrastructural analysis by TEM further substantiated these observations, revealing nuclear deformation, increased cytoplasmic vacuolation, and marked chromatin disorganization. To assess the downstream molecular consequences, qRT-PCR was performed which revealed robust up-regulation of hypertrophic markers (*Nppa*, *Mef2c*, *Myh6*, *Myh7*, *Actc1*, *Ttn*, *Nfatc1*, and *Nfatc2*), with *Nppa* and *Mef2c* showing >12-fold induction in p.R721Q, p.A780V and p.C786Y variants. Additionally, an increased apoptosis marker *Bax/Bcl2* ratio, particularly in p.R721Q (=27.6) and p.A780V (=18.3), confirmed activation of apoptotic signalling pathways. Additionally, 2D and 3D *in-silico* modeling revealed notable conformational changes which strongly corroborate *in-vitro* findings. Collectively, these variants weaken the structure and function of UNC45B, causing disrupted sarcomeric organization, pronounced cellular hypertrophy, and subsequent activation of apoptotic pathways leading to cardiac dysfunction.

## 1 Introduction

Dilated cardiomyopathy (DCM), affecting approximately 1 in 2500 individuals [1–3], is characterized by left ventricular enlargement and impaired systolic function in the absence of abnormal loading conditions [4,5]. To date, mutations in over 100 genes have been linked to DCM [6]. It causes progressive deterioration in cardiac contractility, primarily attributed to disruptions in the mechanisms of force generation and transmission within the sarcomere. While the structural proteins form the foundational framework of the sarcomere, the assembly and maintenance of this intricate contractile apparatus also depend critically on the activity of molecular chaperones. Among these, myosin chaperones are essential for the proper folding and maturation of myosin polypeptides, whose motor domains are structurally complex and incapable of achieving their native conformations autonomously. Through facilitating the correct folding of myosin, these chaperones ensure the functional integrity of sarcomeric architecture and thereby preserve myocardial performance [7].

Members of the UCS (**U**NC-45/**C**RO1/**S**he4P) protein family, particularly Uncoordinated mutant number 45 (UNC45), are indispensable in mediating the folding of the myosin motor domain into a mechanochemically competent state [8]. Despite their critical function, the precise molecular mechanisms underlying the interaction between UNC45 and myosin remain poorly understood. UNC45 plays an essential part in myosin assembly throughout sarcomerogenesis. Originally identified in *C. elegans* (Unc45), presented as two isoforms, namely UNC45A and UNC45B, in mammals. While UNC45A is ubiquitously expressed in all mammalian cells-types, UNC45B is predominantly found in skeletal and cardiac striated muscle cells [9]. UNC45 is structurally organized into three distinct domains, each contributing to its functional versatility. The N-terminal tetratricopeptide repeat (TPR) domain facilitates interactions with the molecular chaperone heat shock protein 90 (HSP90). The central domain, although unique to UNC45, has an as-yet-undetermined function. The C-terminal UCS (**U**NC-45/**C**RO1/**S**he4P) domain is critical for its role in myosin folding and stabilization [10]. Recent studies have highlighted the essential role of UNC45B in stabilizing cardiac myosin, maintaining myofibrillar organization critical for heart contractility, and preventing the development of cardiomyopathies [11]. Deficiency in UNC45 has been linked to DCM and was effectively replicated in experiments involving myosin knockdown, suggesting that UNC45 may serve as a potential therapeutic target for the treatment of cardiomyopathies [11].

In this study, WES of familial and sporadic DCM cases, followed by screening of 100 DCM cases by Sanger sequencing uncovered seven rare *UNC45B* variants. *In-vitro* functional assays showed that mutants displayed elevated protein levels, abnormal cell morphology, and significant up-regulation of hypertrophy and apoptotic markers. Complementary *in-silico* predictions further highlighted structural disruptions, collectively suggesting that these variants impair myofibrillar organization and contractile function through hypertrophic signaling.

## 2. MATERIALS AND METHODS

### 2.1 Enrolment of study subjects and clinical evaluation

The present study involved five families (FDCM1-FDCM5), each comprising at least two affected and one unaffected member, alongside ten sporadic DCM cases (SDCM1–SDCM10), all of whom underwent WES. Additionally, 100 sporadic DCM cases (proband age< 25 years), were enrolled for screening by Sanger sequencing. Further, 100 age-matched healthy controls, lacking clinical evidence of cardiac abnormalities, were also included. Diagnosis was based on left ventricular (LV) dimensions and functions, specifically Left Ventricular Ejection Fraction (LVEF) <45% and LV fractional shortening (LVFS) <25%. Patients with hypertension, coronary artery disease (CAD), or congenital heart disease (CHD) were excluded. The study received approval from the Institutional Ethics Committee.

### 2.2 Genomic DNA isolation and Whole Exome Sequencing (WES)

Isolation of genomic DNA (gDNA) was carried out from peripheral blood samples using the conventional ethanol precipitation method. Five families (FDCM1-5) and 10 sporadic cases (SDCM1-10) were subjected to whole-exome sequencing. WES libraries were prepared by using the Nextera Rapid Capture Expanded Exome Kit (Illumina, USA), according to the manufacturer’s protocol. Sequencing was performed on the Illumina HiSeq2500 platform, producing 2×150 bp paired-end reads per sample with an average coverage of 100X. The resulting datasets underwent comprehensive quality control assessments to ensure data integrity.

### 2.3 Sanger Sequencing

Besides WES analysis, we also performed Sanger sequencing of 100 sporadic DCM patients. Protein-coding regions of *UNC45B* (NM_173167.3) were amplified by Polymerase Chain Reaction (PCR). The PCR-amplified products were enzymatically purified using Exonuclease I (USB Affymetrix, Inc.) and recombinant Shrimp Alkaline Phosphatase (USB Affymetrix, Inc.). They were then directly sequenced using the BigDye Terminator v3.1 Cycle Sequencing Kit (Applied Biosystems, Inc.) on DNA Analyzer (ABI-3130, Applied Biosystems, USA). Sequencing data were analyzed by using FinchTV chromatogram viewer (http://www.geospiza.com/ftvdlinfo.html, Geospiza, Seattle, WA, USA). Besides screening sporadic cases by conventional Sanger sequencing, the identified variants were also screened in other family members to determine their segregation pattern.

### 2.4 NGS data analysis

Analysis of the data was carried out after sequencing had been completed. The sequencing reads were mapped to the human reference genome (GRCh38 or hg19) using alignment algorithms. Base quality score recalibration was performed using the GATK Recalibrator to enhance variant calling accuracy. Finally, variant calling was executed to generate VCF files for each sample. A comprehensive description of the NGS data analysis workflow is available in Supplementary Figure 1 as described in [13].

### 2.5 Prioritization of potentially pathogenic variants

The most important part of data analysis is the prioritization of potentially disease-related variants [13]. Initially, variants with a read depth of ≥20 were short-listed and matched with a curated gene panel. Further filtering was applied using a minor allele frequency (MAF) threshold of <0.01 based on data from the Genome Aggregation Database (gnomAD; https://gnomad.broadinstitute.org/), the 1000 Genomes Project (https://www.internationalgenome.org), and the ExAC database (https://exac.broadinstitute.org/). The shortlisted variants were then classified into distinct categories, including missense, frame-shift (insertions/deletions), nonsense (premature stop codons), synonymous (silent), splice-site, and intronic variants, based on their effect on protein sequences.

### 2.6 *In-Silico* Pathogenicity Prediction

The potential pathogenicity of the detected variants was analysed using VarCards (http://varcards.biols.ac.cn/ ), an integrated database that combines genetic and clinical data for coding variants. VarCards evaluates the functional impact of coding variations through 23 predictive algorithms, including SIFT, PolyPhen2_HDIV, PolyPhen2_HVAR, MutationTaster, MutationAssessor, FATHMM, PROVEAN, CADD, VEST3, M-CAP, REVEL, and FATHMM_MKL.

### 2.7 Conservation analysis of identified variants

To assess the evolutionary conservation of the identified variants, analyses were performed at both the genomic and protein levels. At the nucleotide level, conservation was evaluated using computational tools, viz. GERP, phyloP, phastCons, and SiPhy, which quantify evolutionary constraint across multiple species. Complementarily, protein-level conservation was investigated through cross-species comparison of amino acid sequences using the NCBI HomoloGene database (http://www.ncbi.nlm.nih.gov/homologene).

### 2.8 Prediction of secondary and three-dimensional structures

The impact of the aforementioned genetic variations on protein conformation was assessed by predicting alterations in both secondary and tertiary structures. Secondary structures of the WT and mutant UNC45B protein variants (p.L486F, p.C487Y, p.A492T, p.D496Y, p.R721Q, p.A780V, and p.C786Y) were analyzed using the PDBsum tool (https://www.ebi.ac.uk/thornton-srv/databases/pdbsum/). Additionally, three-dimensional homology modeling of both the WT and mutant proteins was carried out using AlphaFold2 servers to generate their respective structures. Subsequently, the modeled mutant structures were superimposed with WT-UNC45B using UCSF Chimera, and the Root mean square deviation (RMSD) values were calculated.

### 2.9 Protein stability prediction

Several computational tools were utilized to assess the impact of variations on the structural stability and dynamics of the mutated UNC45B proteins. These include DynaMut2 (https://biosig.lab.uq.edu.au/dynamut2/) [14], a tool that combines vibrational entropy and protein flexibility analysis to predict the effects of mutations on protein dynamics and stability. MUpro (https://mupro.proteomics.ics.uci.edu/) [15] provides machine-learning-based predictions of protein stability changes induced by single-site mutations. CUPSAT (https://cupsat.brenda-enzymes.org/) [16] uses environment-specific atom potentials and torsion angle distribution to predict the effects of amino acid substitutions on protein stability. DUET (https://biosig.lab.uq.edu.au/duet/stability) [17] integrates two complementary approaches, mCSM and SDM, to provide reliable predictions of mutation-induced changes in stability. Additionally, I-Mutant (https://folding.biofold.org/i-mutant/i-mutant2.0.html) [18] offers both sequence-based and structure-based predictions for assessing the impact of single-point mutations on protein stability. Collectively, these tools provide valuable insights into the molecular mechanisms underlying the effects of variations on protein structure and function.

### 2.10 *In-vitro* functional analysis

#### 2.10.1 Cloning and Site-Directed Mutagenesis

The *Homo sapiens UNC45B* clone was procured from Genscript (Accession no. NM_173167.3, Clone ID: OHu30162). The full-length coding sequence of *UNC45B* was sub-cloned into the pcDNA3.1/NT-GFP TOPO TA (Invitrogen) vector as per the manufacturer’s protocol. The fidelity of the UNC45B construct was confirmed through Sanger sequencing.

Wild Type (WT)-UNC45B was used as a template for preparing mutant constructs (c.1456C>T, c.1460G>A, c.1474G>A, c.1486G>T, c.2162G>A, c.2339C>T and c.2357G>A) by site-directed mutagenesis using Quick change II XL Site-directed mutagenesis kit (Agilent Technologies Inc., Santa Clara, CA, USA) as per manufacturer’s instructions.

#### 2.10.2 Cell culture and transfection

Rat cardiomyoblast H9c2 cell line (purchased from cell repository, NCCS, India) was used specifically for the generation of stable cell lines. These cells were cultured in Dulbecco’s Modified Eagle’s Medium (DMEM) (Gibco, Life Technologies Corp.) supplemented with 10% Fetal Bovine Serum (Gibco, Life Technologies Corp.) at 37°C in a humid atmosphere with 5% CO2. Cells were seeded at a confluency of ∼10[ cells per well of 6-well tissue culture plates and transfected with UNC45B-WT plasmid 12 hours after seeding, using FuGENE®-6 HD transfection reagent (Promega Corp.). Cells were allowed to recover for 48 hours post-transfection in regular growth media to ensure proper expression of the transgene. A kill curve was prepared to determine the minimal G-418 disulfate (Antibiotic) concentration that kills untransfected cells. Only cells that successfully integrated the plasmid and expressed the antibiotic resistance gene survived, while non-transfected cells died. Once resistant colonies were visible, individual colonies were isolated using limiting dilution and expanded in G418-containing DMEM media (with 10% FBS) followed by validation of stable lines by Real-time PCR. After validating the stable UNC45B-WT cell line, the same protocol was employed to generate UNC45B mutant cell lines (UNC45B-L486F, UNC45B-C487Y, UNC45B-A492T, UNC45B-D496Y, UNC45B-R721Q, UNC45B-A780V, and UNC45B-C786Y).

#### 2.10.3 Western blot

To investigate the impact of UNC45B missense variants on protein expression, western blotting was carried out. UNC45B-stable cell lines (both WT as well as mutants UNC45B-L486F, UNC45B-C487Y, UNC45B-A492T, UNC45B-D496Y, UNC45B-R721Q, UNC45B-A780V, and UNC45B-C786Y) were seeded in 6-well plates and harvested once they reached a confluency of >90% per well. Cell lysates were prepared in RIPA buffer (20 mM Tris-Cl pH =7.0, 250 mM NaCl, 1% NP40, 10% Glycerol, 5 mM EDTA, 20 mM sodium fluoride, 1 mM orthovanadate, 1 mM phenylmethyl sulfonyl fluoride supplemented with 1X protease inhibitor cocktail (Roche, Switzerland)). The lysates were resolved in 8% SDS-polyacrylamide gel and transferred to PVDF membrane (Bio-Rad Laboratories). The membrane was blocked in 3% BSA diluted in TBST (with 0.1% Tween 20) for 1.5 h at room temperature (RT), incubated overnight with UNC45B-specific primary antibody (Abnova, Invitrogen) at 4°C. After incubation with primary antibody, the blot was washed for 4 min (5X) with TBST and then incubated with HRP-conjugated goat anti-mouse IgG antibody (Genei, Merck Specialties Pvt. Ltd.) for 1h at RT. Blot was washed with TBST (5 times, 5 min each) and detected with ECL detection kit (WesternBright ECL-HRP Substrate). Mouse heart tissue lysate was taken as positive control.

#### 2.10.4 Immuno-staining

To investigate cellular localization of WT and mutant UNC45B variants (p.L486F, p.C487Y, p.A492T, p.D496Y, p.R721Q, p.A780V, and p.C786Y), stable H9c2 cell lines expressing these constructs were seeded onto glass coverslips placed in 6-well culture plates. Cultures were incubated overnight at 37°C in a 5% CO[ environment. Once the cells reached 70-80% confluency, they were washed twice with 1X PBS and fixed with 4% paraformaldehyde (PFA) for 10 minutes. Prior to immuno-staining the cells were permeabilized with 0.1% Triton X-100 diluted in 1X PBS for 15 minutes followed by incubation for 2 hours in blocking buffer (1% BSA in PBS) at RT. Subsequently, the cells were incubated with primary antibody (Anti-GFP antibody, Invitrogen, USA, 1:200 dilution) in 0.1% BSA for overnight at 4°C. After thorough washing with 1X PBST (5X, 5 min each), the samples were treated with Alexa Fluor 488-conjugated Goat anti-rabbit IgG (Molecular Probes, OR, USA) for 2 hours at RT. Post-incubation, cells were washed extensively with 1X PBST (5-6X), followed by cytoskeletal staining with Phalloidin (Sigma-Aldrich, MO, USA) and nuclear staining with DAPI. The stained cells were mounted with mounting medium and imaged using the SP8 STED Laser Scanning Super-Resolution Microscope System by Leica. Image analysis was done by using Image J software (NIH, Bethesda, MD, USA).

#### 2.10.5 Transmission electron microscopy (TEM)

Cultured cells were fixed in a freshly prepared fixative solution containing 2.5% glutaraldehyde and 2% paraformaldehyde in 0.1 M sodium phosphate buffer (pH 7.4). The fixation was carried out initially for 20 minutes at room temperature, followed by incubation for 4-6 hours at 4 °C to ensure optimal preservation of cellular ultrastructure. Post-fixation was performed in 1% osmium tetroxide (OsO ) for 1 hour at 4 °C to enhance membrane contrast. Subsequently, the samples were thoroughly washed in buffer, dehydrated through a graded series of acetone, and infiltrated sequentially with toluene and embedding resin. The specimens were finally embedded in Araldite CY212 resin (TAAB Laboratories, UK) and polymerized for sectioning. Semi-thin sections (0.5 µm) were prepared using an ultra-microtome (Leica Ultracut UC7, Austria), mounted on glass slides, and stained with aqueous toluidine blue to assess fixation quality and identify regions of interest for ultrathin sectioning.

For transmission electron microscopy (TEM), ultrathin sections of grey-silver interference color (approximately 60-70 nm thick) were obtained and mounted on 300-mesh copper grids. These sections were subsequently contrasted with 1% aqueous uranyl acetate followed by 0.5% lead citrate (pH 13) for 10 minutes each, with gentle rinsing in distilled water after every step. The grids were examined using a TALOS F200S transmission electron microscope (Thermo Fisher Scientific, Waltham, MA, USA) operated at 200 kV. Digital images were captured at suitable magnifications ranging from 2050× to 5500× using the TIA imaging software integrated with the microscope system.

#### 2.10.6 Quantitative Gene Expression Analysis by qRT-PCR

To further substantiate the findings from immunostaining and TEM, quantitative real-time PCR was performed to check the mRNA expression levels of cardiac hypertrophy markers (*Myh6, Myh7, Actc1,Ttn, Nppa, Nfatc1, Nfatc2,* and *Mef2c*) as well as apoptosis marker genes (*Bax, Bcl2, Casp3,* and *Casp9*) in stably transfected H9c2 lines. Total RNAs were isolated from UNC45B-WT and mutant stable cell lines (UNC45B-L486F, UNC45B-C487Y, UNC45B-A492T, UNC45B-D496Y, UNC45B-R721Q, UNC45B-A780V, and UNC45B-C786Y) by TriZol method as per the manufacturer’s protocol and treated with RNase-free, DNase I (Thermo-Fisher Scientific, USA), and reverse transcribed into cDNA using RevertAid First Strand cDNA Synthesis Kit (Thermo Fisher Scientific). The mRNA expressions of hypertrophic as well as apoptotic marker genes, as mentioned above, were determined using the real-time PCR assay kit KAPA SYBR® FAST qPCR Master Mix Kit (Merck KGaA, Darmstadt, Germany) by QuantStudio5 qPCR System (Applied Biosystems, USA). GAPDH was used as the internal control. The experiments were performed in triplicate, and the data were plotted as fold change with the standard error of mean. The sequences of primers used for amplification are given in Supplementary Table 2.

### 2.11 Prediction of RNA secondary structure

Following the assessment of protein instability, we extended our analysis to evaluate mRNA structure at the transcript level. To achieve this, the target RNA sequence was obtained from the *UNC45B* transcript (NM_173167.3). For this purpose, short RNA segments were designed, each consisting of 41 nucleotides in length. To accurately assess the impact of the variation, the mutation was strategically positioned at the 21^st^ nucleotide, thereby placing it at the centre of the segment that ensured the inclusion of 20 nucleotides flanking the mutation site on either side, providing a comprehensive context for the sequence environment surrounding the variation.

Once the RNA sequences were prepared, the secondary structures of WT and mutant RNA fragments of UNC45B were analyzed using the RNAStructure Web Server (version 6.0.1) [19]. This platform utilizes thermodynamic-based algorithms to predict RNA secondary structures, providing comprehensive insights into folding conformations and base-pairing dynamics within the RNA sequences. Moreover, MutaRNA tool was also employed to perform mutational analysis of RNA fragments, examining the structural modifications induced by each missense variation [20].

### 2.12 Statistical analysis

Statistical analyses were conducted using unpaired Student’s *t*-test to compare differences between WT and experimental groups individually. Data obtained from Western blot and RT-qPCR experiments were expressed as mean fold change ± standard error of the mean (SEM) from three independent experiments. A *p*-value < 0.05 was considered statistically significant.

## 3. Results

### 3.1 Identification of genetic variants in *UNC45B*

WES of 5 families and 10 sporadic cases, as well as Sanger sequencing of an additional 100 unrelated sporadic DCM patients, identified a total of 9 variations in *UNC45B* gene. Out of which, 7 were non-synonymous (c.1456C>T: p.L486F, c.1460G>A:p.C487Y, c.1474G>A:p.A492T, c.1486G>T:p.D496Y, c.2162G>A:p.R721Q, c.2339C>T, p:A780V and c.2357G>A:p.C786Y) and 2 were synonymous variations (c.2307C>T:p.Y769= and c.2340G>T:p.A780=). Out of 7 non-synonymous variations, 4 variations were identified by WES and rest 3 were detected by Sanger sequencing (see Table 1). The Variants identified by WES are as follows: a novel variation c.1456C>T: p.L486F was detected in one of the family (FDCM3), an already known variation c.2339C>T, p: A780V, was observed in another family (FDCM4); two other variations, c.2162G>A: p.R721Q and c.2357G>A: p.C786Y, those were also identified in sporadic DCM cases i.e., (p.R721Q, in SDCM5 & SDM7) and (p.C786Y in SDCM2, SDCM4 and SDCM7). Further, three other variants, namely, c.1460G>A: p.C487Y, c.1474G>A:p.A492T, and c.1486G>T:p.D496Y, were identified by conventional Sanger sequencing by screening of 100 sporadic DCM cases. We reported three novel variations to the Clinical Variation database, and obtained the following accession numbers: *SCV002564410* (c.1456C>T: p.L486F), *SCV005684996 (*c.1460G>A, p.C487Y), and *SCV005685002* (c.1486G>T, p.D496Y). Minor allele frequency (MAF) of the identified variants was checked in both global as well as Indian databases (shown in Table 1). According to the global gnomAD database, MAF of c.1460G>A:p.C487Y and c.1486G>T:p.D496Y variants were not reported (Novel), while the remaining variants had an MAF of <0.001 (except c.2357G>A, p.C786Y (MAF=0.004)) (Table1). The IndiGenomes database, which focuses specifically on genetic variants from the Indian population, showed the majority of variants had allele frequency <0.01, except c.2162G>A:p.R721Q (MAF=0.0375) and c.2357G>A, p.C786Y (MAF=0.07). The pathogenicity scores of each variant were predicted using the VarCards server, revealing that most were categorized as ‘extremely damaging’ (>70% in each case), except for c.1474G>A:p.A492T, which had a score of <70% (Table 1B). Given these findings, we proceeded with all the variants for further downstream analysis.

**Table 1.**
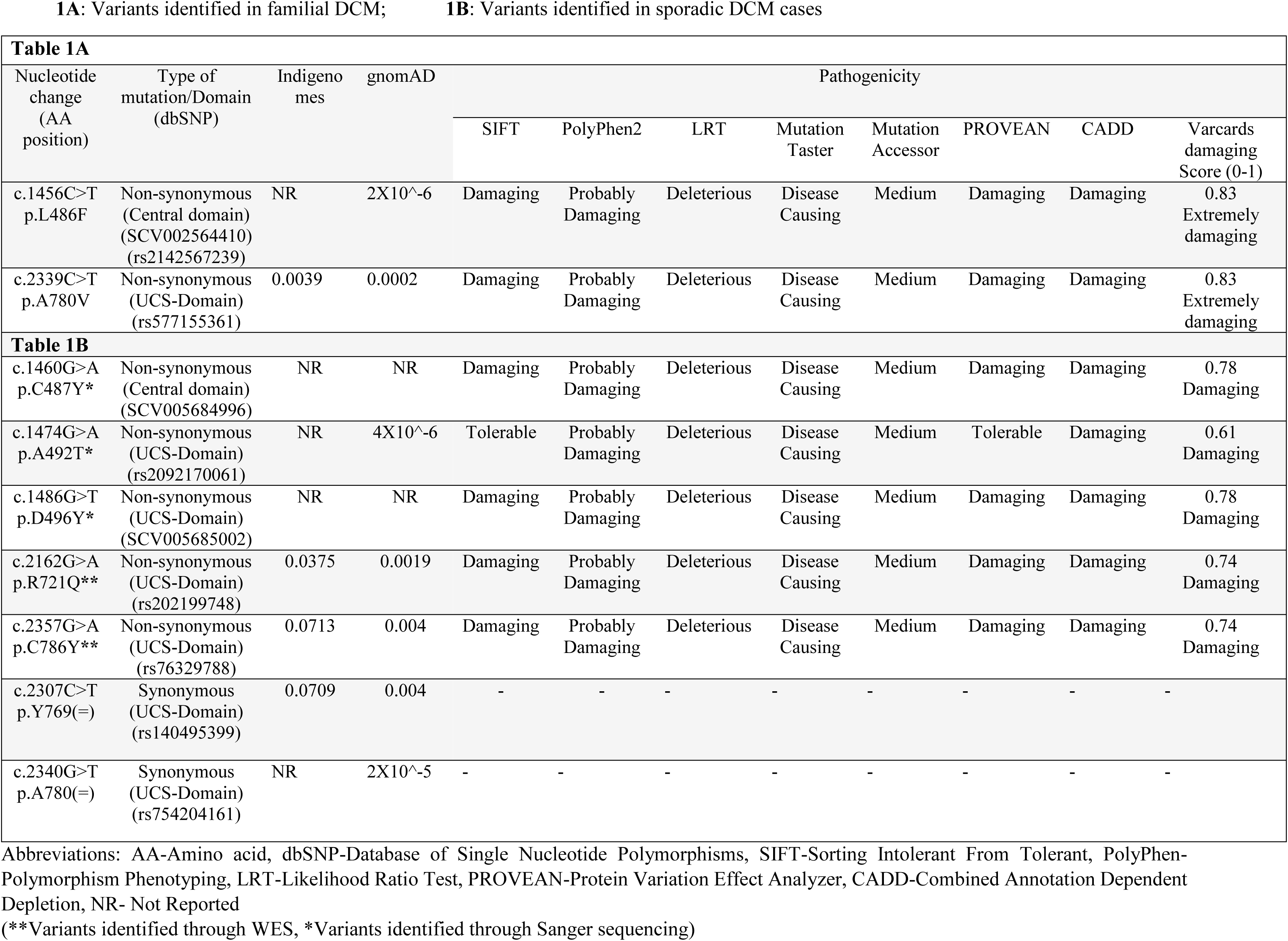
A: Variants identified in familial DCM; Table 1B: Variants identified in sporadic DCM cases

Interestingly, out of 7 non-synonymous variants, 2 variants (p.L486F and p.C487Y) were localized to the Central domain of UNC45B, whereas the rest five variants (p.A492T, p.D496Y, p.R721Q, p.A780V and p.C786Y) were confined to the UCS domain (Figure 2B).

### 3.2 Analysis of segregation pattern of family members by Sanger sequencing

Variants identified through WES were independently validated using Sanger sequencing. In DCM- Family3, the heterozygous UNC45B variant c.1456C>T (p.L486F) was detected in the affected father and elder brother of the proband, while it was absent in the unaffected mother and younger sister. Similarly, in DCM family-4, the c.2339C>T (p.A780V) variant was observed in both the proband and affected father, whereas it was not found in other family members. The corresponding chromatograms from Sanger sequencing, confirming these variants, are presented in Figures 1A and 1B. However, analysis of the available family members from sporadic DCM cases revealed no evidence of inheritance of these mutations, thereby confirming the truly sporadic nature of those cases.

**Figure 1:**
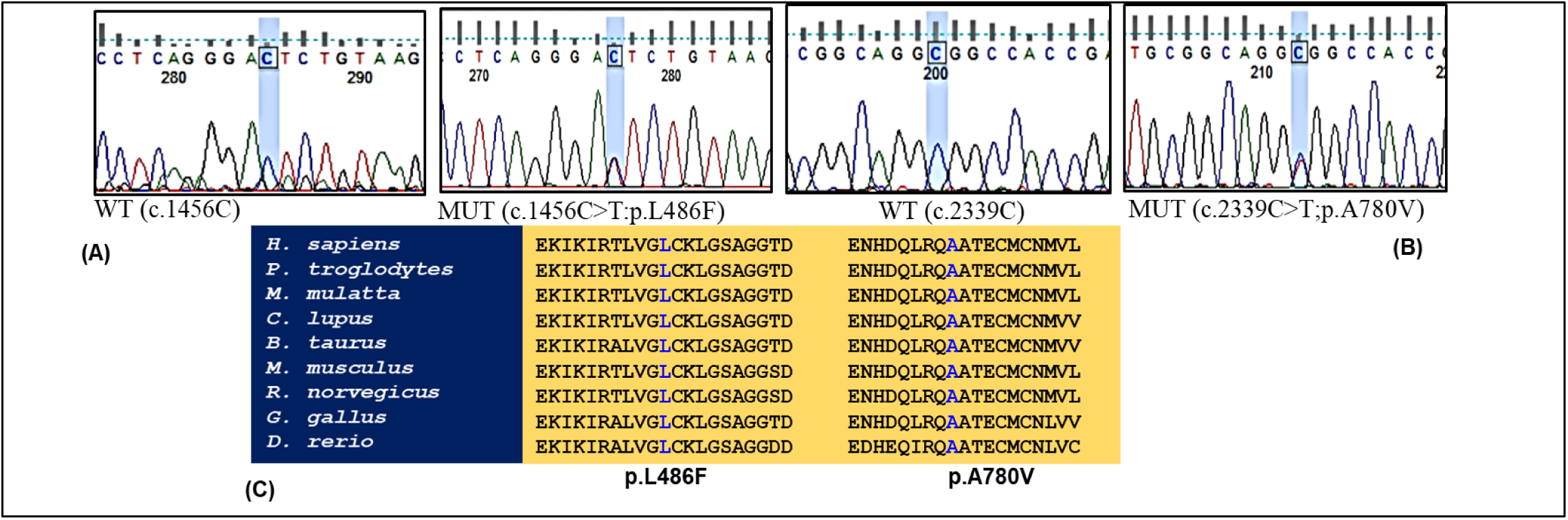
Sequence chromatogram of wild-type and mutant genetic variants. WT, Wild-type; MUT, Mutant; **(A)** DCM family-3, and **(B)** DCM family-4 underwent mutation analysis by Sanger sequencing. **(C)** Evolutionary conservation analysis of genetic variants

**Figure 2:**
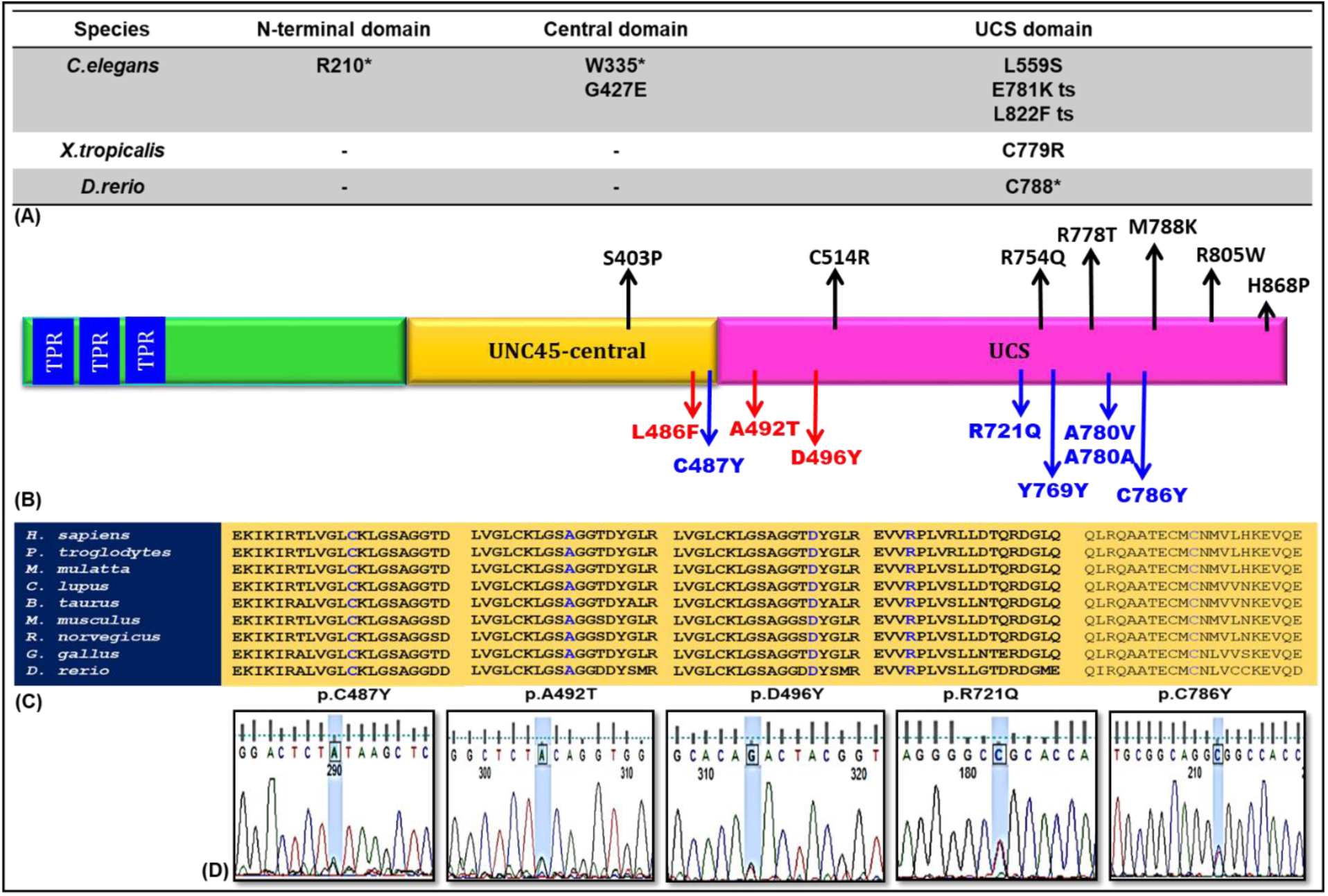
Schematic representation of localization, phylogenetic conservation and sequence chromatogram of missense *UNC45B* variants. **(A)**UNC45B variants identified in other species retrieved from literature; (**B**) The *UNC45B* protein with 931 amino acids showing the N-terminal tertratricopeptide repeat (TPR), Central domain, C-terminal UCS domain. Genetic variants reported in literature are written in black and seven genetic variants identified in our study are written in red/blue colour; **red ones are novel variants**; (**C**) evolutionary conservation of variants across the species. (**D**) Sequence chromatogram of wild-type and mutant genetic variants. WT, Wild-type; MUT, Mutant

### 3.3 Evaluation of cross-species conservation for the detected variants

Sequence comparison of UNC45B proteins across taxa demonstrated that the amino acid sequences for each variant were highly conserved across a wide range of species, spanning from non-mammalian species such as *D. rerio (*NP_705959.1*), G. gallus (*XP_046758531.1) to *R.norvegicus* (NP_001402006.1), *M.musculus* (NP_848795.3), *B.Taurus* (XP_002695676.1), C. lupus familiaris (XP_537726.2), M. mulatta (XP_028692035.1), *P. troglodytes* (XP_016787481.1) and *H. sapiens* (NP_775259.1) (Figure 1C and Figure 2C).

Evolutionary conservation analysis using multiple computational tools such as GERP, phyloP, phastCons, and SiPhy, demonstrated that the identified *UNC45B* variants reside within highly conserved genomic regions. GERP scores, which designate sites under evolutionary constraint with a cut-off value of 2, revealed values exceeding 4.5 for all variants, indicating conserved, non-neutral sites. Similarly, phastCons whose cut off value ranges from 0 to 1 and the same value for SiPhy scores is 12.17, consistently placed these variants within regions of high conservation (Supplementary Table 1).

### 3.4 Effect of missense variants on secondary and tertiary protein structures

Interestingly, secondary structure analysis indicates that the UNC45B protein consists exclusively of α-helices, with no β-sheets or β-helices. A comparison of the predicted secondary structures of WT-UNC45B with its mutant proteins (p.L486F, p.C487Y, p.A492T, p.D496Y, p.R721Q, p.A780V, and p.C786Y) revealed alterations, including either loss or formation of α-helices (highlighted by red circles in Figure 3) as well as changes in coiled regions at various locations across the protein (Figure 3). Additionally, three-dimensional homology modeling of both the WT and mutant proteins was carried out using AlphaFold2 servers to generate their respective structures. Models for both variants were predicted with a confidence score exceeding 90%. Subsequently, the modelled-structures were aligned using UCSF Chimera, and the RMSD values for WT and mutant UNC45B proteins were calculated (Figure 4). The ranking of protein distortion, based on RMSD values in descending order, was as follows: p.R721Q (7.760) **>** p.C786Y (7.237) **>** p.A780V (6.363) **>** p.L486F (5.756) **>** p.A492T (5.240) **>** p.C487Y (3.199) **>** p.D496Y (2.977).

**Figure 3:**
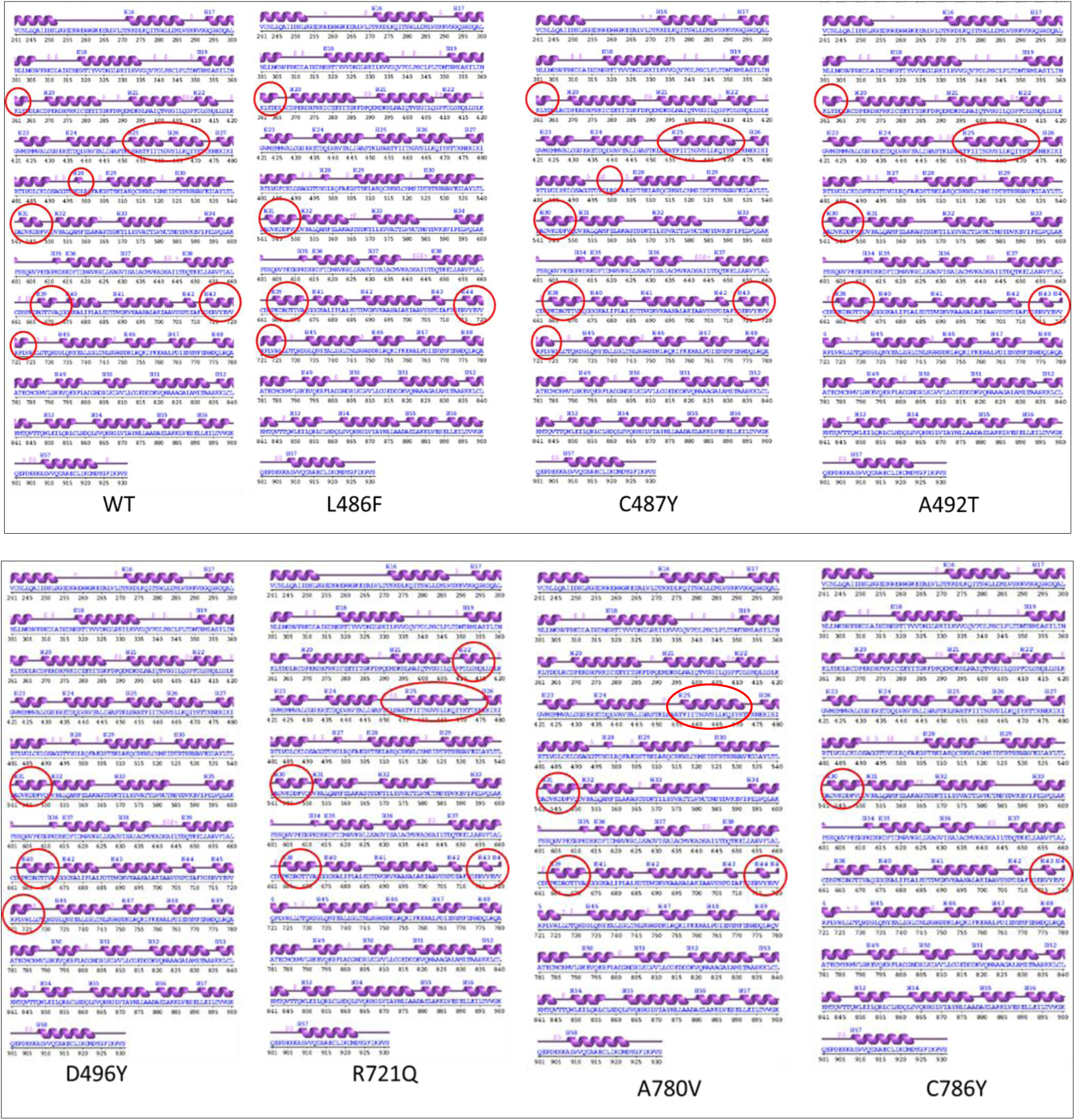
Prediction of the effect of UNC45B missense variants on the secondary structure of mutant proteins; Alpha-helices (H) are represented by purple helical structures whereas β-strands are represented by firm lines.

**Figure 4:**
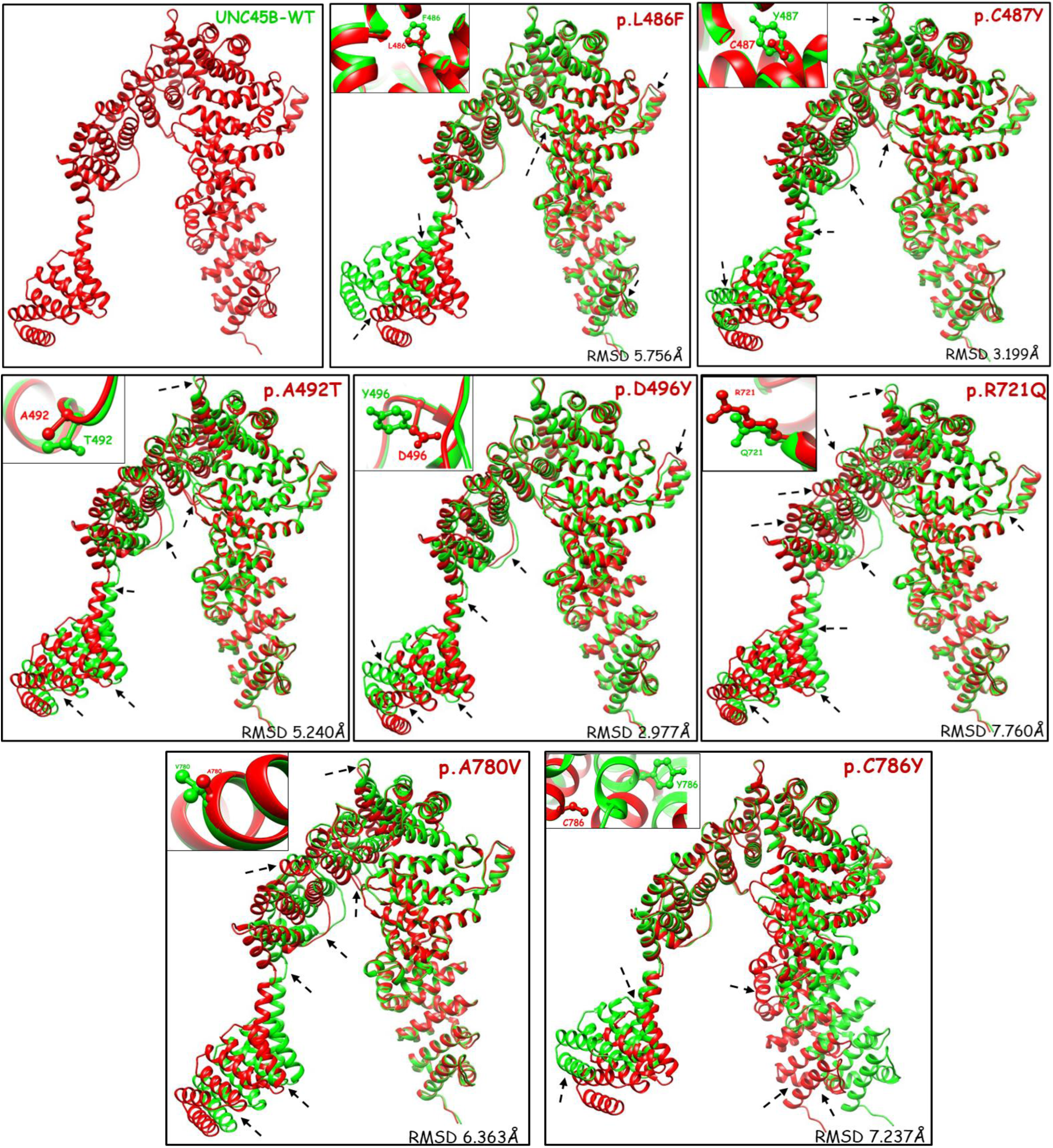
3D model of UNC45B-WT (green), Superimposition of UNC45B-WT (green) with mutant proteins (Red), respectively. WT and substituted residues are shown by ball and stick model (upper right corner). Regions showing differences in superimposition are marked by arrow.

### 3.5 Alterations in protein stability

To further evaluate the impact of the identified mutations on protein stability, we employed a range of *in-silico* prediction tools. Among all variants analyzed, the p.R721Q substitution demonstrated the highest degree of structural destabilization, as consistently indicated across multiple protein stability prediction platforms (Table 2). Notably, this pronounced instability, aligns well with our RMSD analysis, reinforcing the deleterious effect of this specific mutation on the conformational integrity of the protein. In contrast, the remaining variants viz., p.L486F, p.C487Y, p.A492T, p.D496Y, p.A780V, and p.C786Y, exhibited a comparatively moderate yet significant level of destabilization. These findings collectively suggest that while all variants may impair protein function, the p.R721Q variant is likely to exert the most profound structural and functional consequences.

**Table 2:** Analysis of protein stability affected by damaging variants in *UNC45B*

| AA<br>change | DYNAMUT2 | MUpro | CUPSAT | DUET | I-mutant | SDM | mCSM |
| --- | --- | --- | --- | --- | --- | --- | --- |
| p.L486F | -0.45 kcal/mol<br>Destabilizing | -0.9 kcal/mol<br>Destabilizing | Stabilising | -1.239 kcal/mol<br>(Destabilizing) | -0.70<br>(Destabilizing) | -0.69 kcal/mol<br>(Destabilizing) | -1.158 kcal/mol<br>(Destabilizing) |
| p.C487Y | -0.52 kcal/mol<br>Destabilizing | -1.39 kcal/mol<br>Destabilizing | Destabilising | -0.265 kcal/mol<br>(Destabilizing) | 0.41<br>(Stabilizing) | -0.11 kcal/mol<br>(Destabilizing) | -0.432 kcal/mol<br>(Destabilizing) |
| p.A492T | 0.1 kcal/mol<br>Stabilizing | -0.86 kcal/mol<br>Destabilizing | Destabilising | -0.229 kcal/mol<br>(Destabilizing) | -0.54<br>(Destabilizing) | -0.31 kcal/mol<br>(Destabilizing) | -0.522 kcal/mol<br>(Destabilizing) |
| p.D496Y | -0.03 kcal/mol<br>Destabilizing | -0.33 kcal/mol<br>Destabilizing | Stabilising | -0.709 kcal/mol<br>(Destabilizing) | -0.04<br>(Destabilizing) | -1.36 kcal/mol<br>(Destabilizing) | -0.358 kcal/mol<br>(Destabilizing) |
| p.R721Q | -0.55 kcal/mol<br>Destabilizing | -0.75 kcal/mol<br>Destabilizing | Destabilising | -0.077 kcal/mol<br>(Destabilizing) | -1.39<br>(Destabilizing) | -0.41 kcal/mol<br>(Destabilizing) | -0.174 kcal/mol<br>(Destabilizing) |
| p.A780V | -1.06<br>Destabilizing | -0.49 kcal/mol<br>Destabilizing | Stabilising | -0.561 kcal/mol<br>(Destabilizing) | -0.71<br>(Destabilizing) | -1.03 kcal/mol<br>(Destabilizing) | -0.621 kcal/mol<br>(Destabilizing) |
| p.C786Y | -0.54<br>destabilizing | -0.88 kcal/mol<br>Destabilizing | Destabilising | -0.68 kcal/mol<br>(Destabilizing) | 0.21<br>(Stabilizing) | -0.06 kcal/mol<br>(Destabilizing) | -0.904 kcal/mol<br>(Destabilizing) |

Abbreviations: **AA**-Amino acids; **CUPSAT**-(Cologne University Protein Stability Analysis Tool), **SDM**-Site Directed Mutator; **mCSM**-Mutation Cutoff Scanning Matrix;

### 3.6 Impact of the variations on the UNC45B protein expression and cellular localization

To examine whether the identified variants influence UNC45B protein expression, we quantified UNC45B levels in stably transfected H9c2 cardiomyoblast cells. Western blot analysis revealed a distinct primary band at approximately 95 kDa, corresponding to full-length UNC45B, along with two additional lower-molecular-weight bands at around 68 kDa and 58 kDa (Figure 5A). Further, the protein expression pattern demonstrated a consistent elevation in the levels of the full-length UNC45B (∼95 kDa) protein across all mutant constructs when compared to WT. Among these, the p.A780V variant exhibited the highest expression, closely followed by p.R721Q and p.C786Y, showing an increase of 1.49-, 1.48-, and 1.47-fold relative to the WT, respectively (p<0.05). The remaining mutants, p.L486F, p.C487Y, p.A492T, and p.D496Y, also displayed elevated expression, with an average 1.38-fold, suggesting an alteration in their steady-state expression profile. The mean fold-change values ± standard error for each variant are depicted in Figure 5B. Intriguingly, the enhanced intensity of the 95 kDa full-length UNC45B band in mutant constructs appears inversely correlated with their proteolytic cleavage efficiency, implying that mutant proteins undergo less cleavage than the WT. Furthermore, a comparative analysis between the full-length protein and its truncated fragments revealed significantly greater accumulation of cleaved fragments, a hallmark of early apoptotic or stress-related responses, wherein activated caspases cleave target proteins, and the resulting fragments persist longer than the intact form.

**Figure 5:**
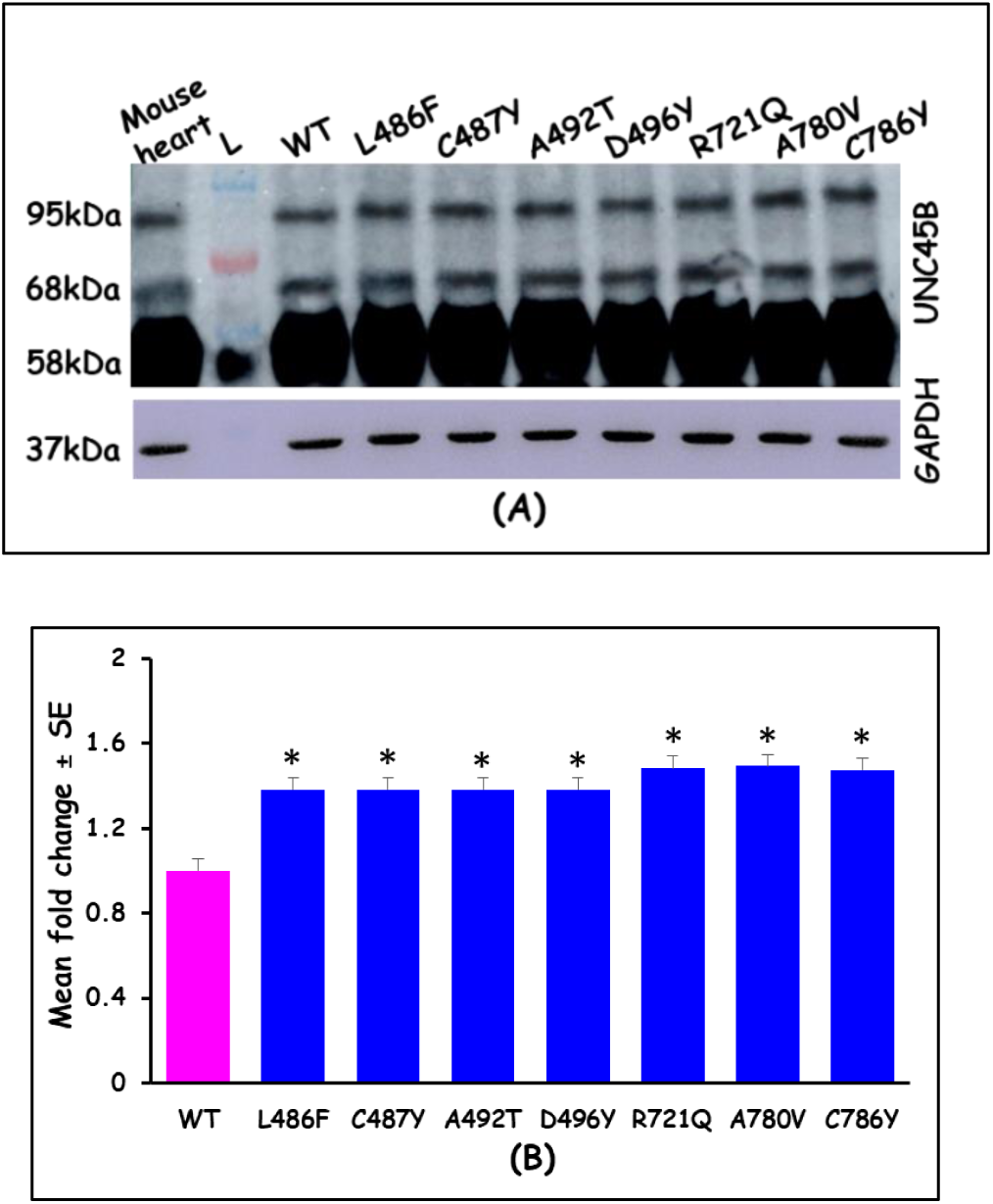
**(A)** Protein expression analysis of UNC45B-WT and mutant proteins by Western Blot; Mouse Heart tissue lysate was taken as positive control**; (B)** The bar chart here represents mean fold change in expression level of full-length (95kDa) WT and mutant UNC45B proteins acquired from three independent experiments (P value <0.05)

The *in-vitro* localization studies in H9c2 cells revealed that both the WT and mutant UNC45B proteins were predominantly found in the cytoplasm. However, significant morphological abnormalities were observed in cells expressing the mutant proteins. Substantial number of cells showed aberrant shape with enlarged size, possibly exhibiting hypertrophic phenotype. Specifically, cells expressing the p.R721Q and p.A780V mutants exhibited enormous increase in size and shape with enlarged nuclei, compared to those expressing the WT protein. These giant cells often displayed fragmented nuclei (Figure 6A). Notably, the frequency of giant cells significantly increased in cells expressing C-terminal UCS domain mutations namely p.R721Q (27.5%), p.A780V (31.25%) and p.C786Y (23.75%) (Figure 6B). These mutant-protein expressing cells display bizarre shapes and sizes, underscores the impact of these mutations on cellular structure and organization.

**Figure 6:**
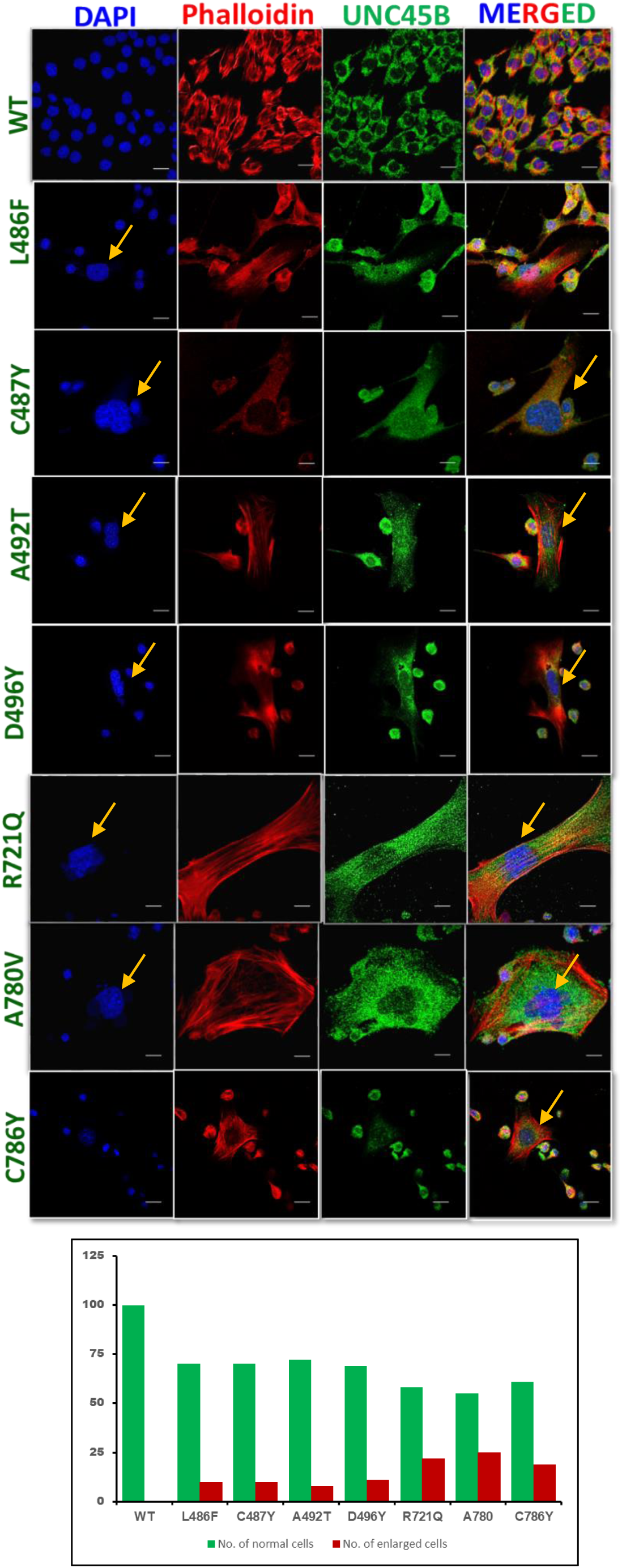
**(A)** Panel **a** (Nuclei: Blue) represents DAPI staining; Panel **b** (F-Actin: Red) represents Phalloidin staining; Panel **c** (UNC45B: Green) represents localization of UNC45B stained in GFP Monoclonal Antibody (3E6); Panel **d** represents a merged image combining DAPI, F-Actin and UNC45B signals. The images were captured at 63X magnification**. (B)** Bar graph showing frequency of giant /hypertrophic cells.

### 3.7 Visualization of cellular structure using electron microscopy (TEM)

TEM analysis revealed a well-preserved ultrastructure and elongated morphology of UNC45B-WT cells. The nucleus (N) appeared clearly defined and centrally positioned, displaying intact dense fibrillar component (DFC), fibrillar center (FC), granular component (GC), and nuclear pore regions. The cytoplasm exhibited uniform electron density, reflecting a homogeneous distribution of organelles, indicative of normal cellular organization and function. In contrast, cells expressing UNC45B mutants (p.R721Q and p.A780V) demonstrated pronounced ultrastructural abnormalities, including cytoplasmic vacuolation, irregularity in nuclear shape and size. The p.R721Q mutant displayed an enlarged cell morphology with a less compact nucleus and dispersed chromatin architecture. The presence of multiple vacuolated regions within the nucleus (indicated by yellow arrows) suggests cellular stress and early degenerative changes. Similarly, the p.A780V mutant exhibited an irregularly shaped nucleus accompanied by numerous electron-lucent cytoplasmic vacuoles (yellow arrows), indicative of extensive cytoplasmic degeneration. The overall loss of cytoplasmic uniformity and increased vacuolization in mutant cells underscore the detrimental impact of UNC45B mutations on cellular integrity.

### 3.8 *UNC45B* variants induce hypertrophic gene expression and activate apoptotic signalling in H9c2 cardiomyocytes

Immunostaining and TEM analysis revealed prominent cellular hypertrophy accompanied by fragmented nuclear morphology in mutant-expressing cells, indicative of an early apoptotic phenotype. To further explore the molecular basis of these observations, we examined the expression profile of hypertrophic markers, including *Myh6, Myh7, Nfatc1, Nfatc2, Nppa*, *Actc1, Ttn, and Mef2c*, using RT-qPCR analysis. A substantial up-regulation of hypertrophic genes was observed across all UNC45B variants when compared with the WT cells. Remarkably, *Nppa* and *Mef2c* exhibited the most pronounced induction among the tested markers. *Nppa* expression was most significantly elevated in A780V and C786Y, demonstrating 22.62-fold and 21.51-fold increases, respectively, followed by R721Q (12.30-fold), D496Y (12.21-fold), A492T (7.68-fold), C487Y (6.88-fold), and L486F (1.14-fold) (Figure 8A). Consistently, *Mef2c* expression was markedly enhanced, with C786Y (19.97-fold), R721Q (19.74-fold), D496Y (16.50-fold), and A780V (13.60-fold) showing the highest induction relative to wild-type (Figure 8B). Similar expression patterns were observed for other hypertrophic regulators and sarcomeric components, including *Nfatc1, Nfatc2, Actc1, Myh6*, *Myh7,* and *Ttn*, each demonstrating significant transcriptional elevation in mutant lines (Figure 8C-H) (Supplementary Table 3).

**Figure 7:**
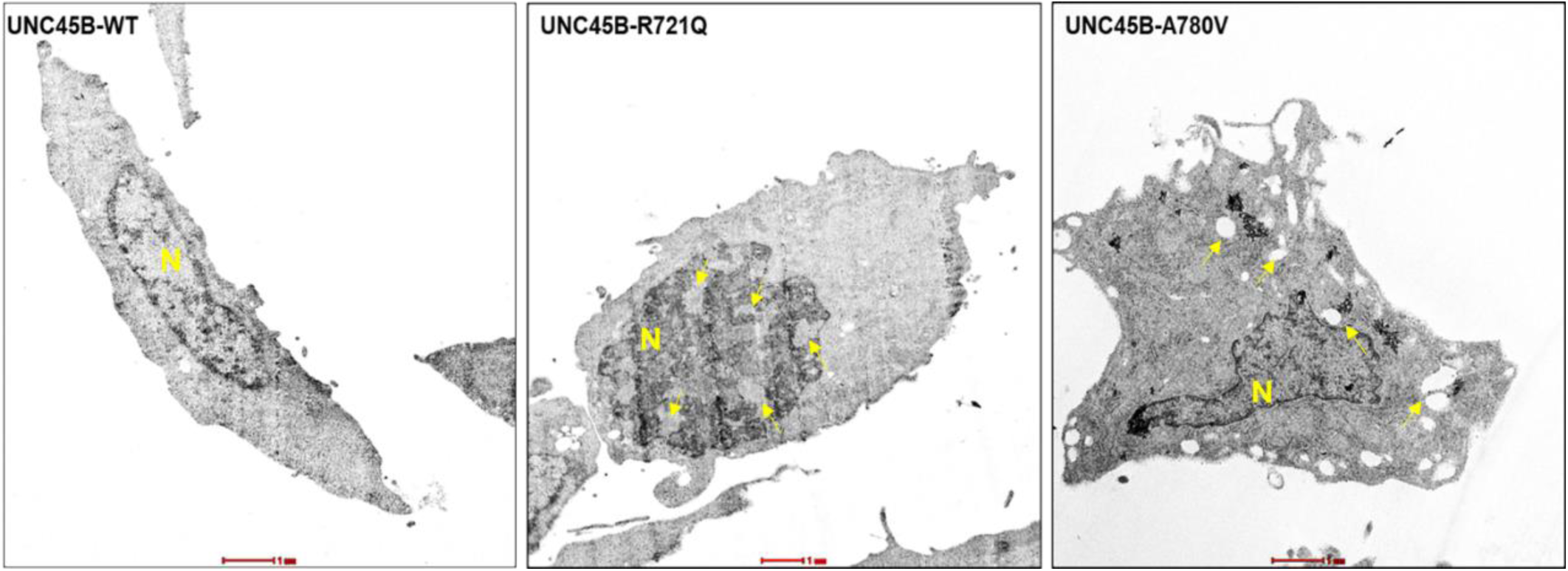
TEM micrographs of UNC45B-WT and UNC45B-mutant H9c2 cells. WT cells exhibit preserved ultrastructure and organized cytoplasm, whereas mutant cells show cytoplasmic vacuoles, disrupted morphology, and nuclear alterations, indicating cellular stress and early apoptotic degeneration (Scale bar=1µm)

**Figure 8:**
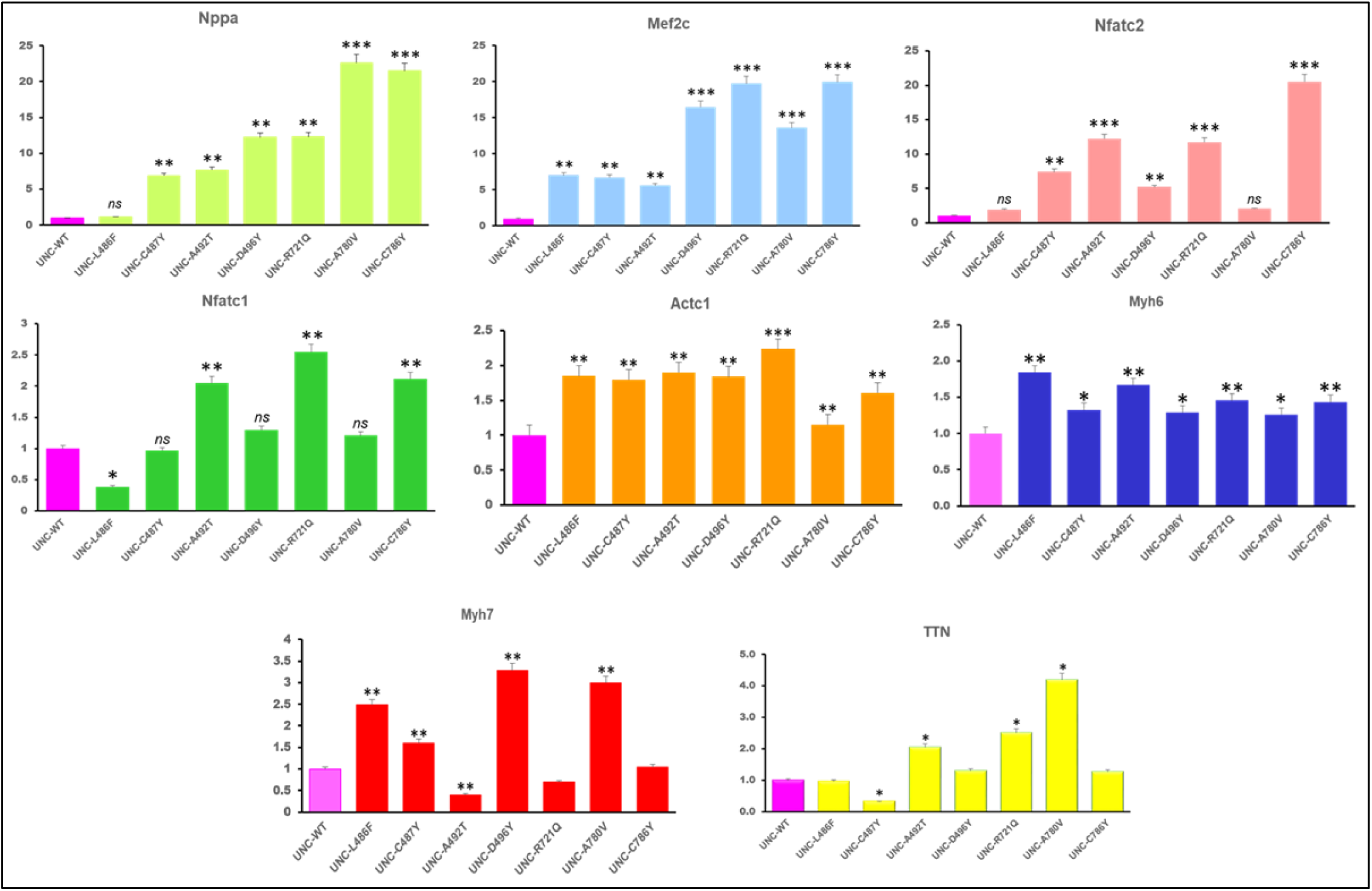
The qPCR detection of the hypertrophic markers. **A-H** The mRNA levels of Nppa, Mef2c, Nfatc1, Nfatc2, Myh6, Myh7, Actc1 and Ttn were determined by qRT-PCR. Three independent experiments for each group. Data are expressed as the mean ± SEM (*P < 0.05, **P<0.01 vs. WT; ***P<0.001 vs. WT)

While hypertrophic growth initially supports cardiomyocyte function, persistent hypertrophy is associated with molecular signalling cascades leading to apoptotic cell death [21]. Therefore, in order to assess apoptotic commitment, we quantified *Bax* and *Bcl2* transcript levels and calculated the *Bax/Bcl-2* ratio. A pronounced increase in apoptotic index was noted in R721Q (27.65) and A780V (18.24) variants, followed by A492T (14.79), D496Y (14.78), C487Y (6.42), C786Y (4.98), and L486F (4.75), suggesting enhanced apoptotic susceptibility associated with UNC45B variants (Figure 9). Collectively, these findings demonstrate that UNC45B pathogenic variants robustly activate hypertrophic signaling pathways and promote apoptosis, reflecting their potential contribution to cardiomyopathic remodeling.

**Figure 9:**
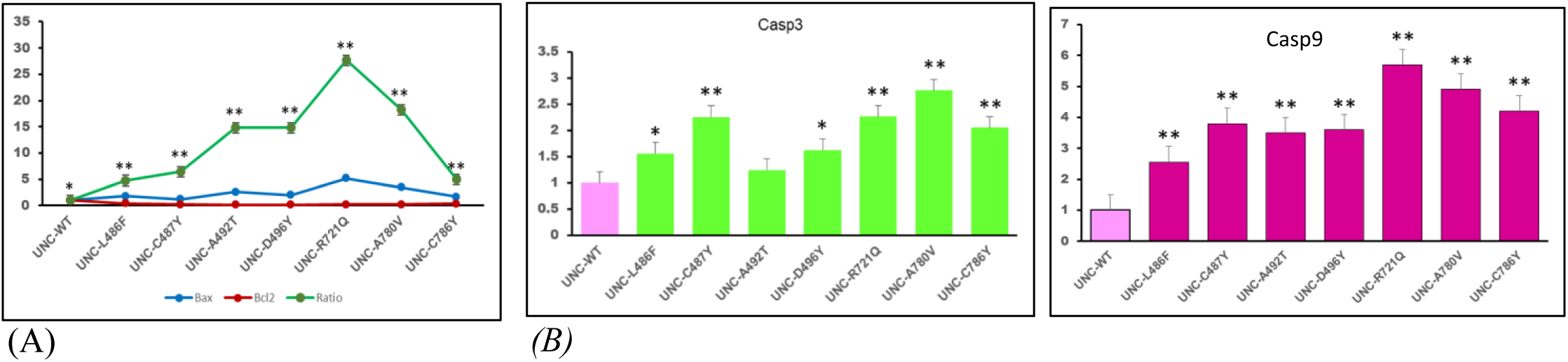
(A) Expression of apoptotic markers (Bax, Bcl2 (**A**) and Casp3 and Casp9 **(B)** by qRT-PCR and estimation of Bax/Bcl2 ratio (apoptotic index)

### 3.9 Alteration of RNA secondary structure

RNA structure is intimately tied to protein structure and function. Any disruption in RNA folding or sequence can lead to errors in protein synthesis, folding, or regulation. Therefore, we further analysed RNA secondary structure that displayed significant disruptions in RNA folding for *UNC45B* variants. The variants c.1456C>T (p.L486F), c.1460G>A (p.C487Y), c.1486G>T (p.D496Y), c.2339C>T (p.A780V), and c.2357G>A (p.C786Y) exhibited pronounced deviations from the WT-RNA secondary structure, suggesting that they might interfere with the normal stability and functional conformation of the RNA molecule. In contrast, c.1474G>A (p.A492T) variant demonstrated relatively minor changes in their RNA secondary structures (Figure 10). Additionally, a quantitative assessment was performed by calculating the relative entropy [H(wt: mu)] of WT and mutant RNA structures (Table 3). The greater the entropy value provided by remuRNA, the more the structural impact of the variant [22,23]. This means, the mutations with higher relative entropy values exhibit greater deviations in RNA structure compared to mutations with lower values. Variant c.2357G>A (p.C786Y) exhibited the highest relative entropy value of 4.593, indicating substantial deviation in RNA structure induced by this mutation, followed by variants c.1486G>T (p.D496Y) and c.1460G>A (p.C487Y) with H(wt:mu) values of 3.577 and 2.629, respectively.

**Figure 10:**
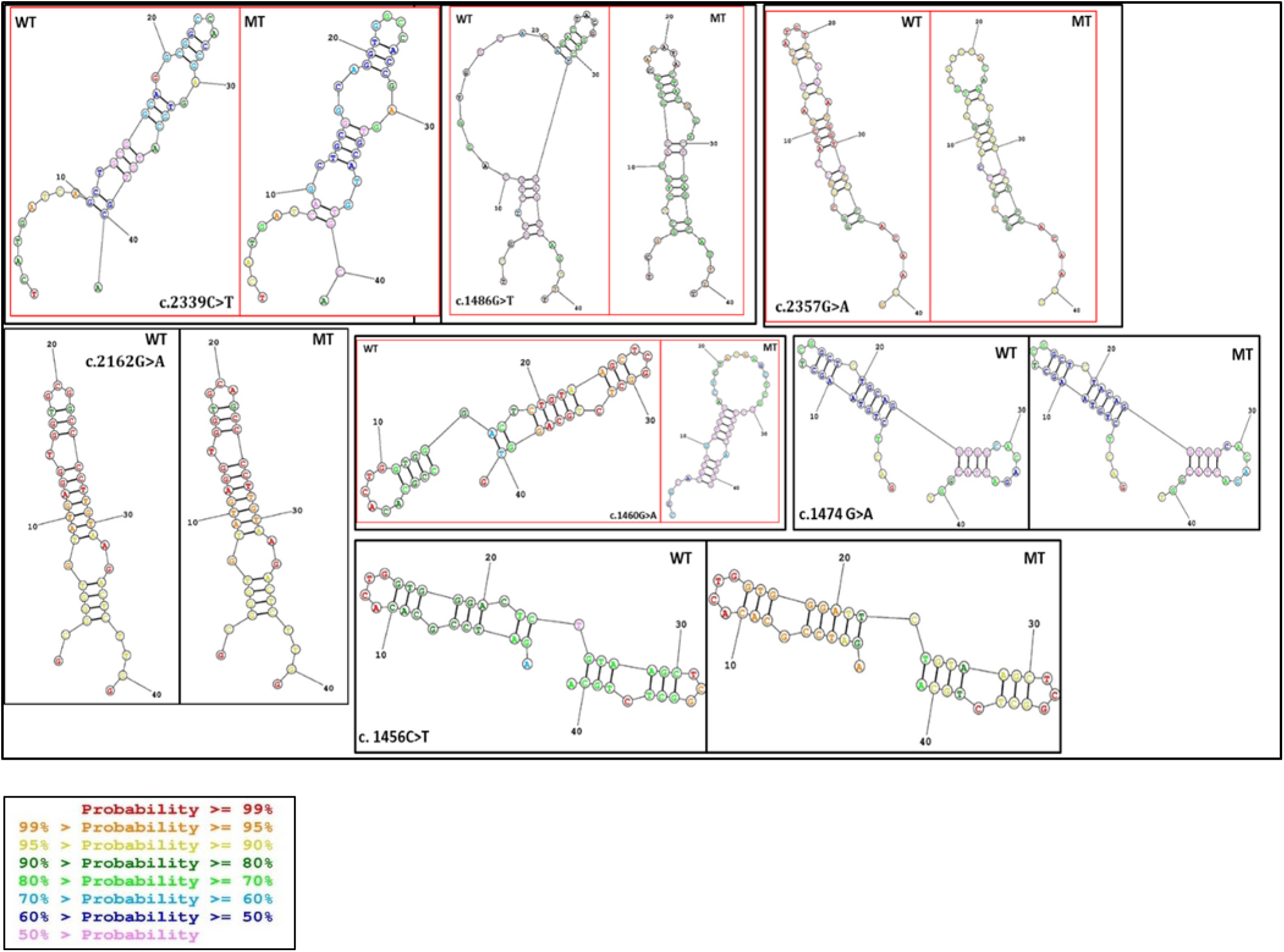
Comparison of Secondary Structures of WT and Mutant (MT) RNA for UNC45B Missense Variants. The WT RNA structure is illustrated on the left, while the mutant RNA structure, altered due to a nucleotide change at the 21st position, is depicted on the right. Base pairing probabilities are represented by varying colors, highlighting structural differences between the two sequences.

**Table 3:** Analyses of the structural impacts of UNC45B missense variants on RNA

| *UNC45B* Variants | H(wt:mu) | MFE(mu) | MFE(wt) | dMFE |
| --- | --- | --- | --- | --- |
| c.1456C>T (p.L486F) | 1.761 | -10 | -8.6 | 1.4 |
| c.1460G>A (p.C487Y) | 2.629 | -7.3 | -9.4 | -2.1 |
| c.1474G>A (p.A492T) | 1.387 | -7.5 | -7.5 | 0 |
| c.1486G>T (p.D496Y) | 3.577 | -6.9 | -7.5 | -0.6 |
| c.2162G>A (p.R721Q) | 0.001 | -11.1 | -11.1 | 0 |
| c.2339C>T (p.A780V) | 1.336 | -7.6 | -9.9 | -2.3 |
| c.2357G>A (p.C786Y) | 4.593 | -7.7 | -10.1 | -2.4 |

Abbreviations: H-Relative entropy between wildtype and mutant RNAs, MFE(mu)-minimum free energy (for the mutant sequence), MFE(wt)-minimum free energy (for the wildtype sequence); dMFE-difference in MFE between mutant and wildtype

Likewise, the effect of the *UNC45* variants on RNA structure is illustrated using multiple graphical approaches, including Circos plots (Figure 11), base pairing probabilities dot plots (Figure 12), differential base pairing probabilities dot plot (Figure 13) and RNA accessibility profile analysis (Figure 14). These MutaRNA findings are related to the effect of variants within the RNA snippet affecting the secondary structure and accessibility to its surrounding context. Prominent deviations from WT patterns were observed in variants p.C487Y, p.A492T, p.D496Y, p.A780V and p.C786Y, indicating significant structural impacts. These base-pair probabilities for WT and mutant (MUT) sequences can also be visualized in heat maps dot matrices (Figure 12). Consistent alterations in base pairing patterns were observed in variants indicating significant structural changes. This is also evident in the entropy chart and the secondary structures predictions using 2D structure and circos plots base pairing probabilities. The differences in base pairing probabilities between mutant and WT RNA (Δ = Pr(bp in WT) - Pr(bp in MUT)) for *UNC45B* missense variants are depicted as differential heat map dot matrices (Figure 13) to compare base pairing probabilities between mutant and WT RNA sequences with red indicating increased interaction likelihood and blue indicating weakened base pairs.

**Figure 11:**
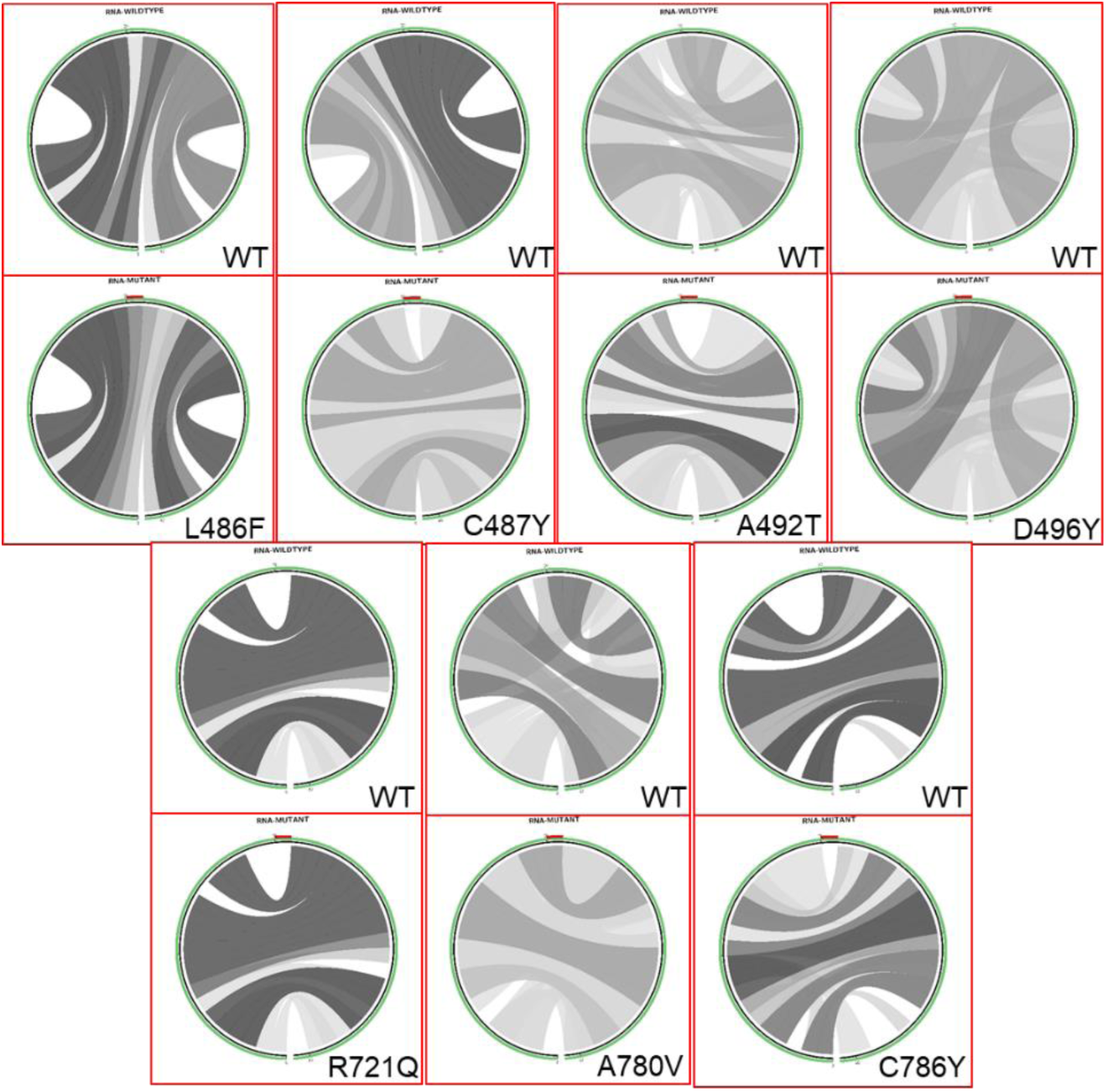
Circos plots illustrate the base pair probabilities for WT and mutant (MT) RNA sequences. The sequences are arranged circularly, starting from the 5’ end at the lower left and proceeding clockwise to the 3’ end. The mutation at position 21 is highlighted in red at the top of the mutant circos plot. Darker shades of gray represent higher base-pairing probabilities

**Figure 12:**
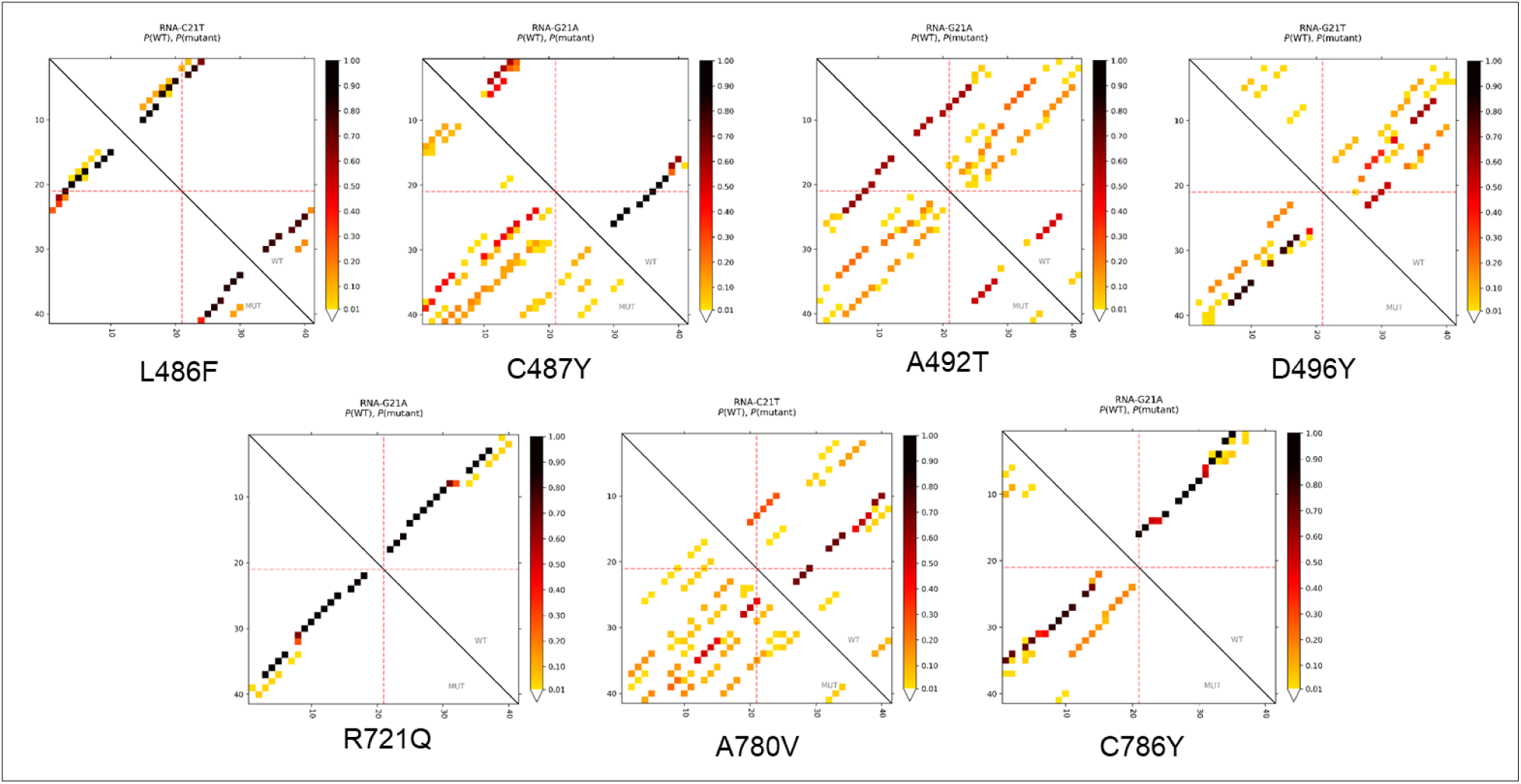
The dot plot illustrates the base-pairing potential for both WT and mutant (MT) RNA variants. The intensity of the dots reflects the likelihood of base pairing between corresponding sequence positions, with darker dots indicating higher probabilities.

**Figure 13:**
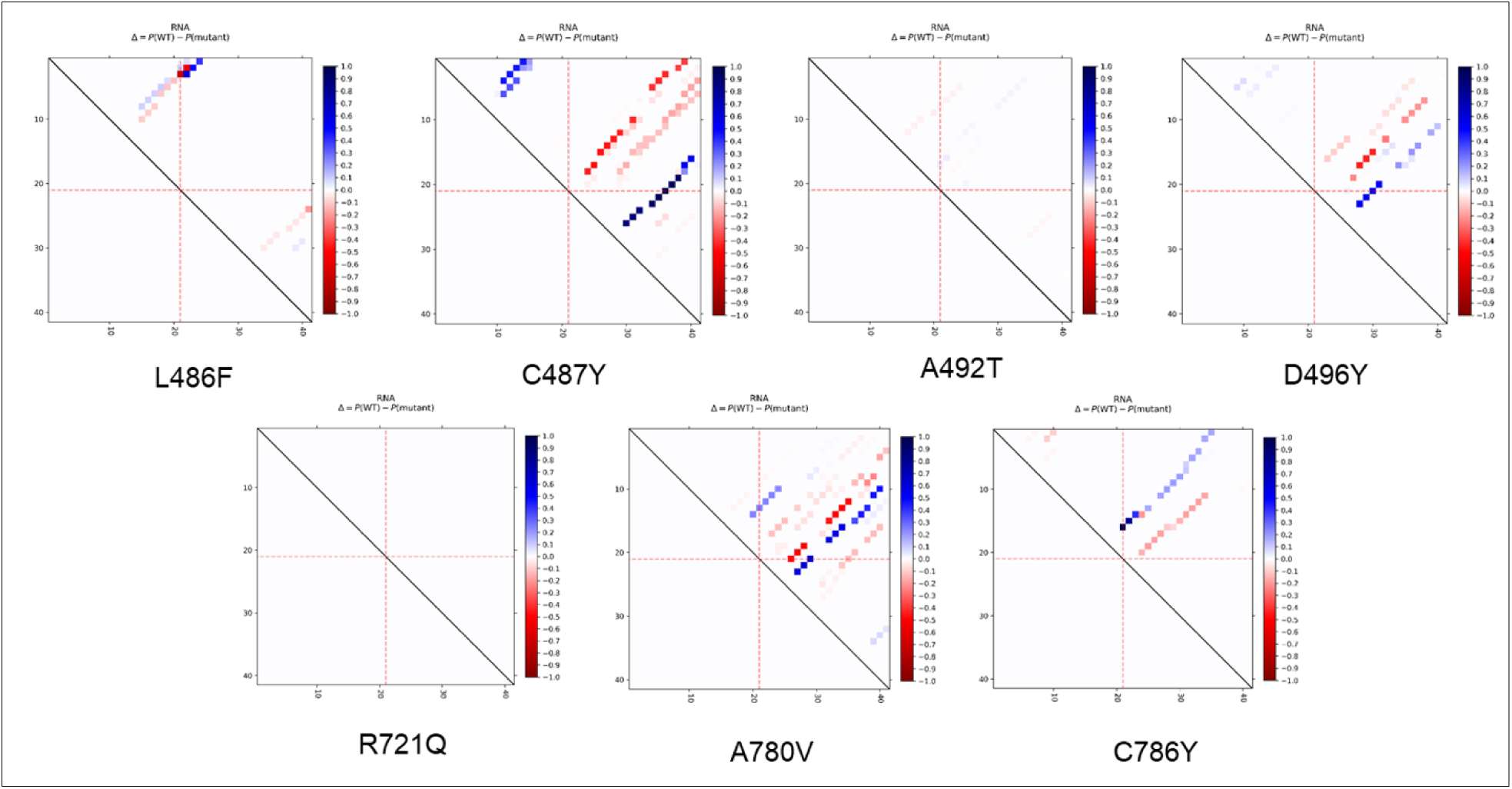
Variation in base pairing probabilities between WT and mutant RNA for UNC45B missense variants. Base pairs weakened by the mutation are shown in blue, while those strengthened are highlighted in red. Axis tick marks indicate intervals of 10 nucleotides, and the mutated position is marked with red dashed lines

**Figure 14:**
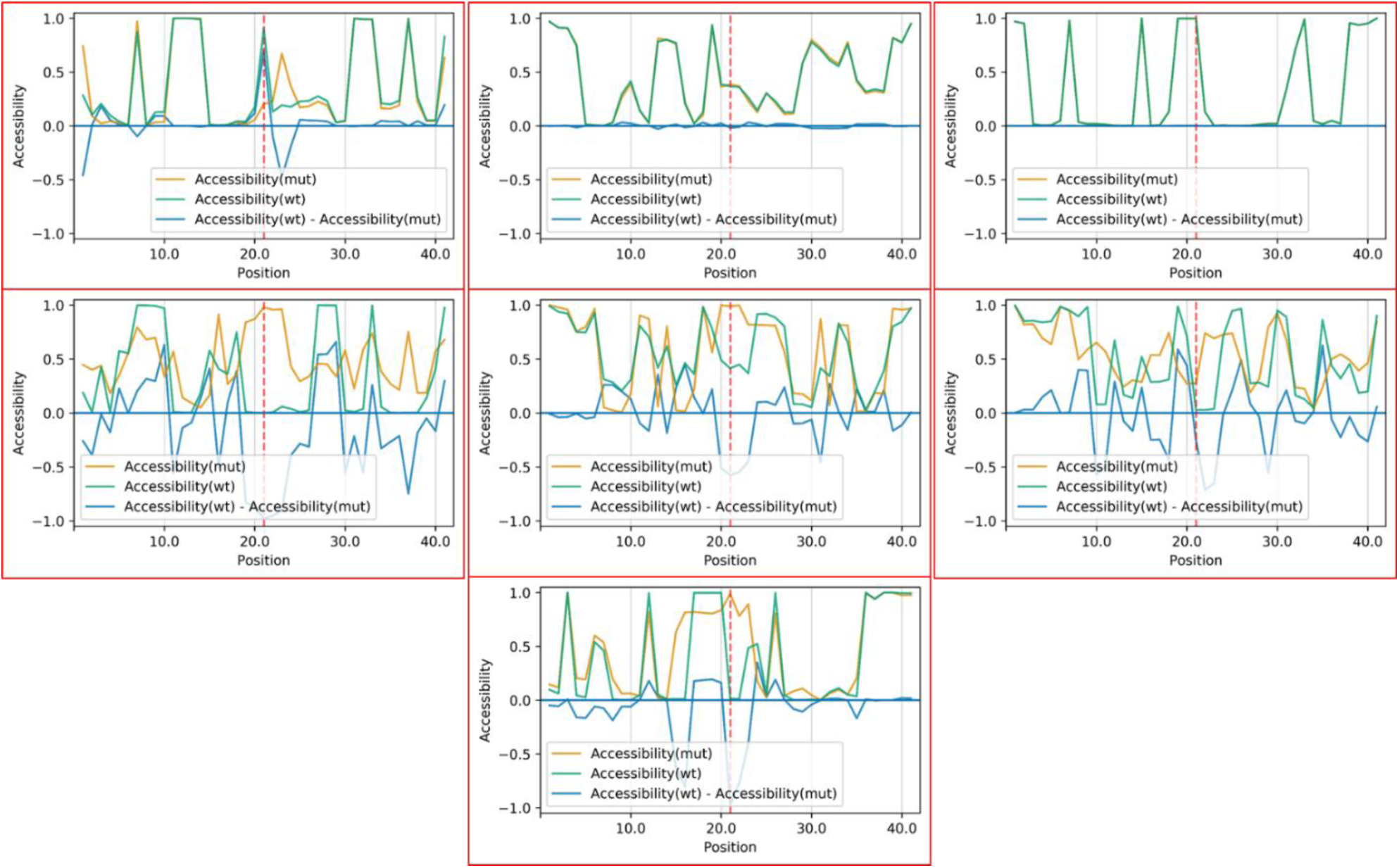
Accessibility profiles of WT and mutant RNA sequences provide insights into how mutations influence single-strandedness, affecting interactions with proteins or other RNAs. Accessibility is assessed as local unpaired probabilities for each nucleotide position. The blue line represents the difference in accessibility (WT-mut), with negative values indicating increased unpaired probability in the mutant. The red line marks the mutated position for each UNC45B missense variant.

### 3.10 Comparative Analysis of RNA Accessibility Profiles in *UNC45B* Variants

Comparison of accessibility profiles of WT and mutant RNA sequences for *UNC45B* missense variants was compared to assess changes in single-strandedness and structure dynamics (Figure 14), which is strongly related to its interactions with other proteins or RNAs. The accessibility profile indicates the likelihood of nucleotides being unpaired for each position within the RNA sequences. The blue line illustrates the alteration in accessibility (WT-mut), where negative values signify positions more likely to be unpaired in the mutant compared to the WT. The “negative drops" in the blue differential accessibility profile signify the reduced accessibility in the folded WT compared to its mutant. The RNA accessibility profiles varied across different variants, with the prominent accessibility differences observed in variants p.L486F, p.D496Y, p.R721Q, p.A780V, and p.C786Y, which further supported RNA secondary structure determination as well as entropy assessment study. These findings suggested that certain variants might significantly impact RNA accessibility, potentially influencing interactions with other molecules or proteins. In contrast, variants such as p.C487Y and p.A492T might have minimal effects on RNA accessibility, highlighting the variability in the structural consequences of the variants. The structural alterations in the predicted secondary structures were also consistent with mutaRNA results.

## Discussion

In the present study, we have reported seven non-synonymous rare variants and two synonymous variants in *UNC45B* gene. Out of seven non-synonymous variants, two (p.L486F and p.C487Y) are located in the central domain, and the remaining five variants, viz., p.A492T, p.D496Y, p.R721Q, p.A780V, and p.C786Y, are situated within the C-terminal UCS domain of UNC45B. Literature survey in model organisms such as *Xenopus*, *C. elegans*, *Zebrafish*, and humans have consistently demonstrated that mutations within the UCS domain (listed in Figure 2) are lethal [24, 25, 26]. In alignment with these findings, our study reveal that 77.7% (7 out of 9) of the variant detected in our cohort are localized within this UCS domain. Structural insights from the crystallographic model of UNC45B in *Drosophila* [27] has shown that the central domain and UCS domain form a contiguous organization of 17 α-helical layers forming five distinct armadillo (ARM) repeat subdomains. Notably, previously reported pathogenic mutations [26] lie within the highest conserved ∼40 amino acid segment that encompasses α-helices (i.e., 18H1-3 and 19H1-2). Interestingly, p.R721Q lies within 17H2 and, p.A780V and p.C786Y in the 18H3 helical regions. Furthermore, specific residue-level interactions, for instance, in *Drosophila*, the Arg803 residue, which corresponds to Arg805 in humans and Arg819 in *C. elegans*, forms a stabilizing inter-repeat ion pair with Glu766 (homologous to Glu767 in humans and Glu781 in *C. elegans*) [27]. Intriguingly, Glu781 in *C. elegans* is known to harbour a temperature-sensitive paralytic mutation, while the corresponding human variant, Arg805Trp, disrupts the same ion-pair interaction. Such alterations are studied to destabilize the α-helical architecture of the UCS domain and compromise its myosin-binding ability. Collectively, these findings strongly support the functional importance of UCS domain. To date, direct evidence implicating *UNC45B* mutations in the pathogenesis of DCM in humans remains limited. However, a biallelic pathogenic variant in *UNC45B* (p.Arg754Gln) has been previously reported in a Lebanese patient affected with congenital myopathy [28]. Furthermore, certain familial cases exhibiting childhood-onset progressive proximal and axial muscle weakness accompanied by respiratory insufficiency have been reported to harbour *UNC45B* variants (c.2332C>T, p.R778W; c.1207T>C, p.S403P; and c.1540T>C, p.C514R), reinforcing the involvement of UNC45B in muscle homeostasis and contractility [29]. Complementary findings in individuals diagnosed with myofibrillar myopathy (MFM) type-II have displayed novel compound heterozygous variants in *UNC45B* (c.2357T>A, p.M786K and c.2591A>C, p.H864P) [30]. However, to the best of our knowledge, our present study constitutes the first clinical and genetic investigation delineating a potential link between *UNC45B* variants and human DCM, thereby broadening the pathogenic landscape of this evolutionarily conserved myosin-chaperone. Nonetheless, UNC45B has been studied in murine models of heart failure; e.g., previous reports have shown that recessive loss-of-function mutations in UNC-45b, particularly in C3H and C57BL/6 inbred mouse strains lead to arrested cardiac morphogenesis, particularly affecting right heart development, and result in severe impairment of myocardial contractility [31].

Although the definite targets for UCS domains are not fully understood, depending on the interacting partner, differential role of UCS domain has been predicted. Through initial bioinformatics analysis using the ‘VarCards’ tool, it is determined that L486F and A780V exhibiting the highest pathogenicity. Notably, these two variants are associated with familial DCM. Conversely, among the seven identified missense variants, A492T demonstrated the least severe impact. Based on pathogenicity scores, the variants can be ranked in decreasing order of severity as follows: L486F & A780V > C487Y & D496Y > R721Q & C786Y > A492T. To gain further insight into the structural consequences of these mutations, we have examined their effects on both the secondary and tertiary structures of the protein. Structural modeling and comparative analyses of the wild-type and mutant proteins reveal notable conformational deviations, as illustrated in Figures 3 and 4. The predicted secondary structure alterations, coupled with the superimposed three-dimensional structural models, suggest substantial disruptions in the spatial organization of the mutant proteins, thereby indicating a potential destabilizing effect [32, 33]. All seven UNC45B variants are identified within evolutionarily conserved regions, underscoring the functional importance of these amino acid residues in preserving the structural integrity and biological activity of the protein. The high degree of conservation at both the nucleotide and amino acid levels suggests strong selective pressure to maintain these sequences across species, reflecting their critical role in proper protein folding and function. To evaluate the potential impact of these mutations on protein stability, a comprehensive computational analyses are performed using multiple ΔΔG (Gibbs free energy change)-based prediction tools, including mCSM, DUET, SDM, MUpro, and CUPSAT. These tools integrate structural, energetic, and evolutionary parameters to estimate the destabilizing or stabilizing effects of each variant. The convergence of predictions from these independent platforms provides a robust assessment of how individual mutations may perturb the native conformation of UNC45B, potentially affecting its functional role in myosin folding and sarcomere assembly [34].

*In-vitro* functional characterization revealed that wild-type UNC45B predominantly localizes within the cytoplasm, indicating its primary site of activity under normal cellular conditions. Previous studies in zebrafish have demonstrated that Unc45b and Hsp90a co-localize with myosin during myofibrillogenesis and associate with the Z line once myofibril assembly is complete. Upon myofiber stress or damage, these proteins dissociate from the Z line and transiently re-associate with myosin [35]. Moreover, UNC45 proteins and their UCS-domain homologues have been shown to facilitate the functional maturation of target myosins across diverse organisms, including *C. elegans*, *Drosophila*, *Saccharomyces cerevisiae*, *Schizosaccharomyces pombe*, zebrafish, and cultured mouse myocytes [36–39]. Immunohistochemical evidence also confirms cytoplasmic localization of UNC45b, UNC45a, and myosin in wild-type embryonic heart tissues [31]. In our study, cells expressing the p.R721Q and p.A780V variants of UNC45B exhibited pronounced morphological alterations, characterized by enlarged cell size and irregular nuclear architecture. Notably, the p.A780V mutant displayed a markedly expanded nucleus and overall hypertrophic phenotype compared to wild-type. These morphological anomalies, accompanied by fragmented nuclei observed through immunostaining, suggest that the p.R721Q and p.A780V mutations trigger a hypertrophic response likely through disruption of coordinated myosin interactions [40, 41]. Such impairment may perturb nuclear-cytoplasmic signaling mechanisms essential for maintaining cellular homeostasis and structural integrity. Pathological cardiac hypertrophy typically arises as an adaptive response to increased workload; however, sustained stress results in metabolic overload, ATP depletion, oxidative stress, and endoplasmic reticulum (ER) stress, thereby activating apoptotic pathways [42]. Chronic activation of these stress pathways induces maladaptive signaling through p53, Bax, and caspase-3 activation, hallmarks of apoptosis. Progressive cardiomyocyte loss through apoptosis subsequently leads to fibrosis, contractile dysfunction, and ventricular dilation, ultimately driving the transition from compensated hypertrophy to heart failure. In this context, hypertrophy and apoptosis act as interconnected signaling processes rather than mutually exclusive events [43]. Hypertrophic stimuli influence apoptotic signaling at multiple levels, contributing to pathological ventricular remodeling. Nuclear fragmentation observed in our mutant lines reflects the terminal phase of apoptosis, involving chromatin condensation, DNA fragmentation, and nuclear disassembly. Mechanistically, this process proceeds via the mitochondrial (intrinsic) apoptotic pathway, where cardiac stress induces mitochondrial outer membrane permeabilization and release of cytochrome c. Cytochrome c associates with Apaf-1 and procaspase-9 to form the apoptosome, which sequentially activates caspase-9 and caspase-3. Activated caspase-3 cleaves ICAD, releasing CAD, which translocates to the nucleus to fragment chromosomal DNA into nucleosomal units, leading to chromatin condensation and nuclear fragmentation [44]. Corroborating this mechanism, our study revealed an increased Bax/Bcl-2 ratio and elevated expression of caspase-3 and caspase-9 in mutant cells compared to wild-type, reinforcing that the p.R721Q and p.A780V variants of UNC45B induce apoptotic signaling secondary to pathological hypertrophy.

UNC45b mRNA has been reported to be highly expressed only in heart and skeletal muscle of adult mice and in the developing hearts of E8.5 C57BL/6 mouse embryos [31]. Its expression at the protein level in H9c2 cell-line has not been reported previously. In this study using H9c2, both immunoblot and immuno-staining experiments show the expression of UNC45B protein in figure 5 and figure 6 respectively. The cleavage sites and proteolytic stability of the UCS domain within UNC45B have been extensively characterized in previous studies. Notably, a detailed structural analysis has been performed by isolating the UCS domain of human UNC45B, followed by pulse trypsin digestion and mass spectrometry. This approach has revealed that the major proteolytic events predominantly occurred within the flexible loop regions situated between armadillo (ARM) repeats, particularly between ARM12 and ARM13. These findings highlight the modular architecture of the UCS domain, wherein the N-terminal half exhibit a compact and protease-resistant structure, while the C-terminal half display increased conformational flexibility and is more vulnerable to proteolytic cleavage. The presence of such structurally dynamic regions underscores the loop segments as primary cleavage hotspots within the UCS domain [45]. Moreover, bioinformatics analysis predicting potential cleavage sites in UNC45B identified two major positions, at residues 529 and 624 (Supplementary figure 2) Cleavage at these sites is expected to generate protein fragments with approximate molecular weights of 58kDa, 68kDa, and 44kDa, suggesting that partial proteolysis of UNC45B can give rise to distinct truncated forms. Consistent with these predictions, our western blot analysis displayed a prominent band at ∼95 kDa, accompanied by additional lower molecular weight bands at ∼68 kDa and ∼58 kDa. These observations suggest that proteolytic processing within flexible loop regions of UNC45B gives rise to truncated yet structurally stable fragments of the protein.

The major function of cardiac myosins is to produce cardiac contraction in order to circulate the blood. The primary objective of this study was to evaluate whether the identified UNC45B variants are associated with phenotypic characteristic of DCM. It is noteworthy that one of our identified novel variants, substitution of leucine at position 486 (p.L486), has previously been implicated in a mouse model. Specifically, at the corresponding position (UNC45b^L486R/L486R) in the C3H mouse strain was reported to result in a significantly reduced cardiac contraction rate in embryonic hearts, approximately half that of wild-type counterparts, indicating a partial loss of UNC45b function [30]. Interestingly, the patient in our cohort harboring the p.L486F mutation exhibited a markedly reduced LVEF of <35%, which is a clinical hallmark of impaired cardiac function in DCM. These findings demonstrate a striking concordance between our human data and previously published murine phenotypes, thereby supporting the potential pathogenic role of the p.L486 position in UNC45B.

Further to examine the impact of missense variants in *UNC45B* on RNA secondary structure, different computational tools have been employed. The prediction of RNA secondary structure implicated RNA folding and stability are influenced significantly by the mutants, suggesting potential functional repercussions at the molecular level. The structural differences between WT and mutant RNAs are quantified through relative entropy calculations, assessing the extent of mutation-induced changes. To validate these findings, multiple visualization techniques are employed, including Circos plots (Figure 11), base pair probability dot plots (Figure 12), and differential base-pairing probability dot plots (Figure 13). Additionally, accessibility profiles, represented as the probability of individual nucleotides being unpaired, were analyzed for both WT and mutant RNA sequences. The differences in accessibility profiles offered insights into how mutations might alter RNA interactions with proteins or other RNAs. These results reveal the wide-ranging structural consequences of the studied UNC45B missense variants. Variants inducing pronounced structural changes in RNA could serve as potential drug targets, paving the way for precision medicine approaches [45]. Furthermore, even minor structural disruptions may influence RNA dynamics and function [46]. These subtle changes can disrupt RNA folding, modify splicing regulatory elements, affect translation efficiency, or act as disease modifiers by compounding the effects of other mutations, potentially leading to a more severe phenotype [47]. Therefore the *in-vitro* analyses aligns with our *in-silico* evaluation strongly suggesting potential functional impact of these variants in DCM pathophysiology.

## Conclusion

In a nutshell, the findings from both *in-vitro* and *in-silico* experiments provide compelling evidence for the critical role of UNC45B in maintaining the cytoskeletal integrity and structural stability of cardiomyocytes. The observed disruptions in cytoskeletal organization in mutants strongly implicate UNC45B as a pivotal regulator of these processes. *In-vitro* studies demonstrated significant aberrations in cellular morphology, including enlarged nuclei, multinucleation, and irregular overall cellular structures in mutant UNC45B. These morphological defects align with impaired actomyosin interactions and compromised cytoskeletal scaffolding, both of which are vital for the mechanical and functional properties of cardiomyocytes. Additionally, *in-silico* analyses, including molecular modeling studies, highlighted the structural consequences of the conformation of mutant UNC45B and its interactions with myosin and associated cytoskeletal components. These disruptions likely impair chaperone activity, critical for the folding and stability of myosin heavy chains. Alterations in binding affinities and interaction dynamics further emphasize the mutation-induced perturbations in cytoskeletal protein networks. These findings open avenues for further research into therapeutic interventions targeting UNC45B-related pathways to restore cytoskeletal integrity and prevent or mitigate the progression of cardiomyopathy.

## Data Availability

All data produced in the present work are contained in the manuscript

## Acknowledgment

We deeply appreciate the patients and their families for their invaluable contributions to this study. We are also profoundly thankful to Padma Shri Prof. T.K. Lahiri and Prof. D. Agrawal from the Department of Cardiothoracic & Vascular Surgery, for their constant encouragement and valuable support in enrolling patients for the study. We are thankful to Sophisticated Analytical Instrument Facility (SAIF), AIIMS, New Delhi for TEM facility and Sophisticated Analytical & Technical Help Institute (SATHI), Banaras Hindu University, Varanasi for Laser scanning super resolution microscopy system. We gratefully acknowledge the Department of Biotechnology, Ministry of Science & Technology, Government of India, for funding this project (BT/PR12369/MED/12/678/2014), and the Indian Council of Medical Research (ICMR) for awarding a Senior Research Fellowship [3/1/1(23)/2022-NCD-I] to Ms. Amrita Mukhopadhyay.

## Author contributions

Bhagyalaxmi Mohapatra: Conceptualization, Supervision, Investigation, Validation, Resources, Writing-Review & editing, Amrita Mukhopadhyay: Data curation, Validation, Software, Formal analysis, Visualization, Writing- Original draft preparation; Ashok Kumar and Dharmendra Jain: for identifying and enrolling patients for the study.

