## Supplementary data for "Whole exome sequencing reveals UNC45B as a novel candidate gene functionally associated with Dilated Cardiomyopathy"

(Supplementary) Table 1: Genomic Conservation Analysis Scores

| Nucleotide Change (AA Position) | **GERP** | | **phyloP** | | **PhastCons** | | **SiPhy** | |
|---|---|---|---|---|---|---|---|---|
| | **Score (cut-off=2)** | **Prediction** | **Score (cut-off>0)** | **Prediction** | **Score (Range 0-1)** | **Prediction** | **Score (cut-off>12.17)** | **Prediction** |
| c.1456C>T (p.L486F) | 4.89 | Conserved | 4.862 | Conserved | 1 | Conserved | 17.57 | Conserved |
| c.1460G>A (p.C487Y) | 4.89 | Conserved | 9.775 | Conserved | 1 | Conserved | 17.57 | Conserved |
| c.1474G>A (p.A492T) | 4.89 | Conserved | 9.775 | Conserved | 1 | Conserved | 17.57 | Conserved |
| c.1486G>T (p.D496Y) | 4.89 | Conserved | 9.775 | Conserved | 1 | Conserved | 17.57 | Conserved |
| c.2162G>A (p.R721Q) | 5.3 | Conserved | 9.614 | Conserved | 1 | Conserved | 18.128 | Conserved |
| c.2339C>T (p.A780V) | 5.2 | Conserved | 7.804 | Conserved | 1 | Conserved | 17.734 | Conserved |
| c.2357G>A (p.C786Y) | 5.2 | Conserved | 9.869 | Conserved | 1 | Conserved | 17.734 | Conserved |

Abbreviations: **GERP**-Genomic Evolutionary Rate Profiling, **phyloP-** Phylogenetic P-value, **PhastCons-** Phylogenetic Analysis with Space/Time Models Conservation Score, **SiPhy-** Si(∑) Phylogenetic Conservation

Supplemnetray Table 2: List of primers used for qRT-PCR

| Genes | Primers |
|---|---|
| *Mef2c* | **FP**-TCAGTCAGTTGGGAGCTTGC |
| | **RP**- TATCTCGAAGGGGTGGTGGTAC |
| *Nppa* | **FP**- GCTTCCAGGCCATATTGGAG |
| | **RP**-GGGGGCATGACCTCATCTT |
| *Actc1* | **FP**- CTGGATTCTGGCGATGGTGTA |
| | **RP**- CGGACAATTTCACGTTCAGCA |
| *Myh6* | **FP**-GCCCAGTACCTCCGAAAGTC |
| | **RP**-GTCCTCCTTTATGGTCACCGTC |
| *Myh7* | **FP**-CTGTCAACACTAAGAGGGTC |
| | **RP**-TTGATCTTCCAGGGTACCC |
| *Ttn* | **FP**-GCTCACCTGTGATCCAGGTC |
| | **RP**- AGGAAGGCGGCTCTTTGATC |
| *Nfatc1* | **FP**-GCCTTTTGCGAGCAGTATCTG |
| | **RP**-GCTGCACCTCGATCCGAAG |
| *Nfatc2* | **FP**- CCACCACGAGCTATGAGAAG |
| | **RP**-GTTTCGGAGCTTCAGGATGC |
| *Bax* | **FP-**GAAGCTGAGCGAGTGTCT |
| | **RP-**TGGCAAAGTAGAAAAGGGCG |
| *Bcl2* | **FP-** TCGCCCTGTGGATGACTGA |
| | **RP-**CAGAGACAGCCAGGAGAAATCA |
| *Casp3* | **FP-**ATGGAAGCGAATCAATGGAC |
| | **RP-**AATGTTTCCCTGAGGTTTGC |
| *Casp9* | **FP-**CACTGGCTCCAACATCGAC |
| | **RP-**AGCCGTGAGAGAGAATGACC |

Supplementary table 3: Mean fold changes in Hypertrophy markers

| *Actc1* | Fold change | Myh7 | Fold change |
|---|---|---|---|
| UNC-WT | 1 | UNC-WT | 1 |
| UNC-L486F | 1.85 | UNC-L486F | 2.490 |
| UNC-C487Y | 1.798 | UNC-C487Y | 1.608 |
| UNC-A492T | 1.9 | UNC-A492T | 0.407 |
| UNC-D496Y | 1.84 | UNC-D496Y | 3.285 |
| UNC-R721Q | 2.235 | UNC-R721Q | 0.700 |
| UNC-A780V | 1.154 | UNC-A780V | 2.991 |
| UNC-C786Y | 1.605 | UNC-C786Y | 1.050 |
| *Nppa* | Fold change | Myh6 | Fold change |
| UNC-WT | 1 | UNC-WT | 1.000 |
| UNC-L486F | 1.142 | UNC-L486F | 1.847 |
| UNC-C487Y | 6.882 | UNC-C487Y | 1.326 |
| UNC-A492T | 7.684 | UNC-A492T | 1.672 |
| UNC-D496Y | 12.21 | UNC-D496Y | 1.291 |
| UNC-R721Q | 12.304 | UNC-R721Q | 1.457 |
| UNC-A780V | 22.62 | UNC-A780V | 1.261 |
| UNC-C786Y | 21.51 | UNC-C786Y | 1.436 |
| *Nfatc2* | Fold change | Ttn | Fold change |
| UNC-WT | 1 | UNC-WT | 1 |
| UNC-L486F | 1.897 | UNC-L486F | 0.97 |
| UNC-C487Y | 7.430 | UNC-C487Y | 0.322 |
| UNC-A492T | 12.243 | UNC-A492T | 2.05 |
| UNC-D496Y | 5.220 | UNC-D496Y | 1.3 |
| UNC-R721Q | 11.760 | UNC-R721Q | 2.51 |
| UNC-A780V | 2.047 | UNC-A780V | 4.19 |
| UNC-C786Y | 20.530 | UNC-C786Y | 1.27 |
| *Nfatc1* | Fold change | | |
| UNC-WT | 1 | | |
| UNC-L486F | 0.3884 | | |
| UNC-C487Y | 0.97 | | |
| UNC-A492T | 2.05 | | |
| UNC-D496Y | 1.3 | | |
| UNC-R721Q | 2.545243864 | | |
| UNC-A780V | 1.21 | | |
| UNC-C786Y | 2.12 | | |
| *Mef2c* | Fold change | | |
| UNC-WT | 1 | | |
| UNC-L486F | 7.04 | | |
| UNC-C487Y | 6.722 | | |
| UNC-A492T | 5.61 | | |
| UNC-D496Y | 16.5 | | |
| UNC-R721Q | 19.74 | | |
| UNC-A780V | 13.6 | | |
| UNC-C786Y | 19.97 | | |

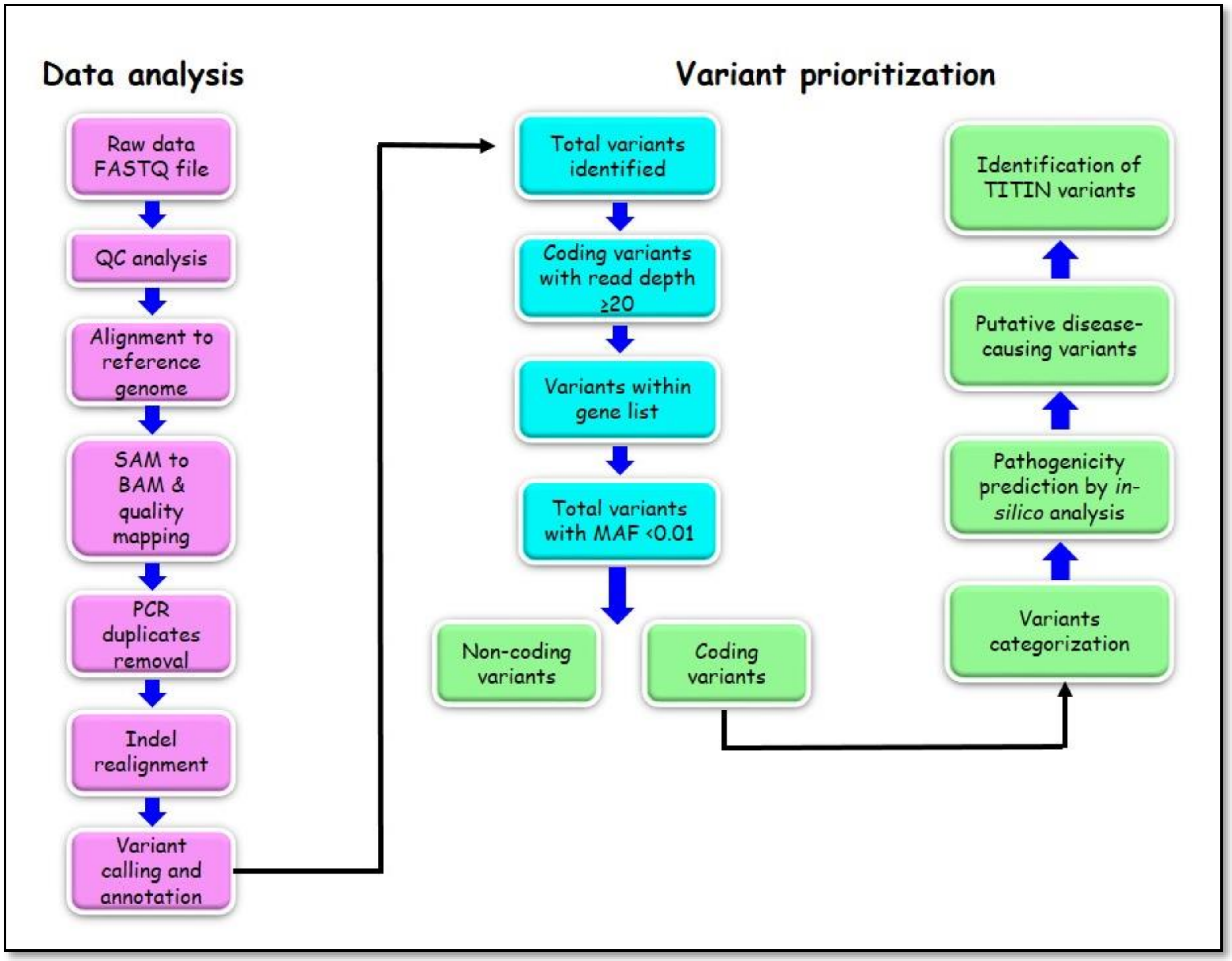


*Supplementary figure 1: WES pipeline for data analysis and variant prioritization*

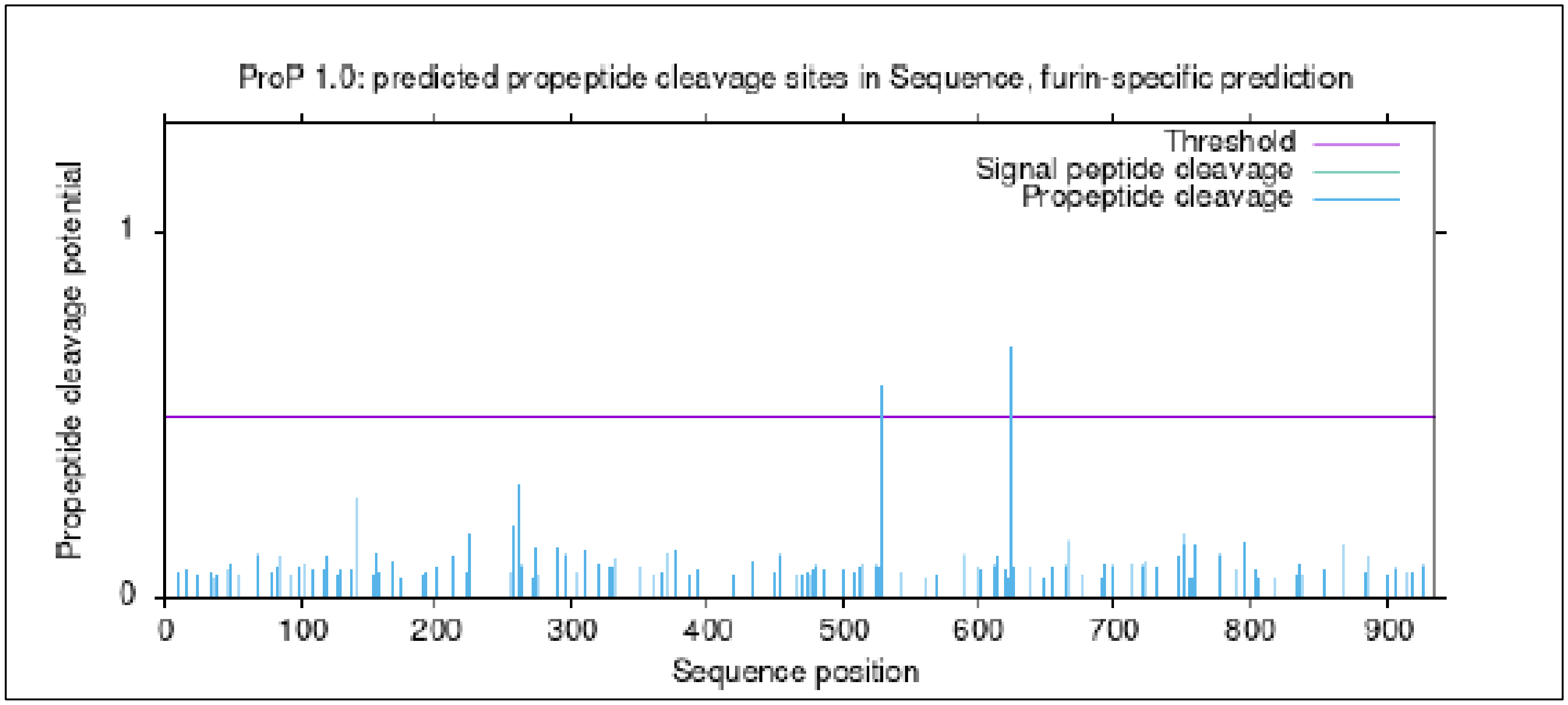

Supplementary figure 2: *In-silico* prediction of proteolytic cleavage sites of UNC45B protein
